# Flexible and efficient count-distribution and mixed-model methods for eQTL mapping with quasar

**DOI:** 10.1101/2025.07.17.25331702

**Authors:** Jeffrey M. Pullin, Jarny Choi, Davis J. McCarthy, Chris Wallace

## Abstract

Identifying genetic variants that affect gene expression, expression quantitative trait loci (eQTLs), is a major focus of modern genomics. Today, various methods exist for eQTL mapping, each using different statistical and methodological approaches. However, it is unclear which approaches lead to better performance, and challenges, particularly scalability as datasets continue to increase in size, remain. Here, we introduce quasar, a flexible and efficient C++ software program for eQTL mapping. Compared to existing eQTL mapping methods, quasar implements a wider variety of statistical models, including the linear model, Poisson and negative binomial generalised linear models, linear mixed model and Poisson and negative binomial generalised linear mixed models. Methodologically, we introduce and implement a simple, analytic approximation to the score test variance in mixed models. Furthermore, we highlight that difficulties with accurately estimating the negative binomial dispersion parameter, previously identified in the context of RNA-seq differential expression analysis, also apply to eQTL mapping. Therefore, quasar implements the Cox-Reid adjusted profile likelihood which enables unbiased estimation of the negative binomial dispersion parameter. We assess quasar’s performance and compare it to three existing eQTL mapping methods: apex, jaxQTL and tensorQTL, on the OneK1K dataset. We demonstrate that quasar’s output agrees with established methods where their models aligns but that quasar is at least 30% and up to 25 times faster. We exploit the range of models implemented in quasar to compare statistical models for eQTL mapping without confounding by implementation. We find that: count-based models have higher power, mixed models do not show better performance in a dataset without substantial relatedness, and the adjusted profile likelihood improves Type 1 error control when using the negative binomial distribution. Additionally, we investigate the relative performance of Poisson and negative binomial mixed models and the use of different approaches for gene-level FDR control. Overall, quasar provides a performant and versatile program for eQTL mapping and we nominate the negative binomial GLM model, incorporating adjusted profile likelihood dispersion estimation, as the statistical model with the best performance.

## Introduction

A major focus of modern genomics is understanding how genetic variation affects gene expression. In particular, statistical methods have been used to identify genetic variants associated with the expression of genes: expression quantitative trait loci (eQTLs) [1]. Traditionally, gene expression has been measured using microarrays or RNA-seq in bulk samples, but the cell-type specificity of expression has motivated a recent shift to using single-cell RNA-seq [2, 3, 4, 5, 6, 7]. Datasets of single-cell eQTLs are rapidly growing in size with the recent releases of the OneK1K dataset, including just under 1,000 individuals, and Phase 1 of the TenK10K project, which aims to map single-cell eQTLs in 10,000 individuals [6, 8]. A major use of eQTLs is linking variants identified by GWAS to affect disease risk to their causal gene and context, for example using colocalisation analysis [9]. Currently, however, there is still a substantial ‘colocalisation gap’, where many GWAS variants cannot be associated with eQTLs, indicating a need for more extensive eQTL discovery across additional contexts and at increased sample sizes [10, 11]. As such, there is an urgent need for scalable eQTL mapping methods that make optimal methodological choices to maximise the power while controlling false discoveries.

Today, while many methods exist for eQTL mapping, there are substantial differences in their methodologies and little empirical comparison of these methodological choices has been performed. A significant difference between methods are the different statistical models of gene expression they use. Earlier methods, such as FastQTL and tensorQTL, modelled transformed expression values using the normal distribution [12, 13]. More recently, methods have modelled the counts produced by RNA-seq with count distribution models, particularly the Poisson and negative binomial distributions, often demonstrating more power compared to methods that use a normal distribution [14, 15]. Use of count distributions has allowed some methods, particularly SAIGE-QTL, to model count expression at single-cell resolution. Furthermore, despite the success of count distribution methods, eQTL methods do not incorporate methodological improvements developed for fitting count models to RNA-seq data for differential expression testing and it is unclear whether these improvements could enhance eQTL mapping performance [16, 17].

Another difference between methods is whether they use mixed models, which include random effects modelling relatedness between individuals and, less frequently, other sources of unwanted variation. Methods exist that implement linear mixed models (LMM) (apex, limix) and Poisson generalised linear mixed model (SAIGE-QTL), in each case incorporating a random effect with covariance matrix set to an estimated genetic relatedness matrix [18, 19]. This approach has been used extensively in the GWAS literature, for example by SAIGE and BOLT-LMM, and is designed to account for cryptic relatedness between samples [20, 21]. However, given the smaller size of eQTL datasets it is unclear whether using mixed models to model relatedness is needed. In addition, it is unclear how different count distributions perform when used as part of a mixed model. Recently, the Poisson generalised linear mixed model (GLMM) has been found to give the equivalent results to a negative binomial GLMM, while while being much faster, when used for single-cell eQTL mapping [5]. However, this finding contradicts extensive work on the importance of accounting for overdispersion using the negative binomial when modelling RNA-seq data [16, 17].

Finally, an ongoing challenge for QTL mapping methods is scalability, especially as datasets increase in size. A bottleneck in cis-eQTL mapping has been the approach used to calculate the significance of genes, where computationally intensive permutations are used to evaluate the significance of the minimum per-gene p-value, followed by gene-level false discovery rate correction using q-values [12, 22]. More recently, the apex and SAIGE-QTL methods have instead used the aggregated Cauchy association test (ACAT) to summarise significance by aggregating the variant-level p-values for each gene [23, 24]. This approach does not require permutations, massively speeding up computation. However, how ACAT interacts with the use of q-values, which have previously been used to control false discovery rates in cis-eQTL mapping, has not been fully described [12, 22].

Here, we introduce quasar, a software package for bulk and pseudobulk eQTL mapping. Compared to existing methods quasar has several advantages. First, it implements a wide variety of methods: linear model, Poisson GLM, negative binomial GLM, linear mixed model, Poisson GLMM and negative binomial GLMM. This wide range of models allows analysts to choose the most appropriate model for their data and allows us, in this paper, to systematically compare different models without potentially confounding inter-package differences. Second, quasar is highly performant and scalable to large datasets, using the ACAT approach to compute gene-level significance, and implements a novel, faster variance approximation to score test variance in mixed models. Finally, quasar implements the Cox-Reid adjusted profile likelihood for dispersion estimation in the negative binomial model [17, 25]. We compare the performance of quasar to three existing methods for eQTL mapping - tensorQTL, jaxQTL and apex - on the OneK1K dataset. Exploiting quasar’s range of implemented models we also compare different approaches for eQTL mapping and study the interactions between methodological choices.

## Results

### Overview of quasar methods

The quasar software package is a command line program, implemented in C++, using the Eigen library for performant linear algebra operations [26] and supporting the common plink bed/bim/fam format for genotype data. The package is freely available from from GitHub (see Code Availability). The quasar software supports both cis- and trans-QTL mapping. Like other QTL mapping methods, quasar uses a score test framework to test for associations between variants and features. Briefly, in this framework, a statistical regression model is first fit for each feature to model and correct for both nuisance covariates, such as age, sex, genotype PCs and gene expression PCs, and, if a mixed model is used, correlation between individual’s measurements. Then, a score test is performed to test for association between genotype and phenotype, using the residualised feature values, for each variant and each feature. In cis-eQTL mapping, the overall significance of genes is commonly assessed. In quasar, the ACAT method is used to combine variant-level p-values to determine significance at the gene level. (See Methods and Supplementary Methods for details)

In addition to the range of models it implements, quasar improves on existing eQTL mapping methodologies in two key ways. First, a key challenge in fitting mixed models using score tests in both QTL and GWAS applications is approximating the variance of the score test. Naively computing the score test variance for each variant requires a *O*(*n*^2^) matrix multiplication, where *n* is the number of individuals, which is too computationally intensive to perform for each variant when *n* is large. Therefore, methods, including SAIGE and SAIGE-QTL, use a variance-ratio-based approximation, which was first developed in GRAMMAR-Gamma (see Methods for details) [14, 21, 27]. Here, we derive and implement in quasar a simple analytic trace-based approximation, which we also show is closely related to the residual-variance-based variance approximation used in the regenie software package for GWAS analysis [28]. Second, while recent methods have used the negative binomial model for eQTL mapping, it is not well understood whether the improved estimation methods developed for negative binomial models in the context of RNA-seq differential expression analysis may be needed [15]. We implement in quasar the Cox-Reid adjusted profile likelihood (APL), which allows unbiased estimation of the dispersion parameter in the negative binominal model [17, 25]. The use of the APL is a core feature of the statistical methodology of the edgeR and DESeq2 packages for differential expression analysis in RNA-seq data [17, 29]. We explore whether this methodology can improve Type 1 error control in the context of eQTL mapping.

### Method validation

To assess the performance of quasar, we applied it to the recent large OneK1K dataset with singlecell RNA-seq of just under 1000 individuals [6]. Briefly, we performed standard filtering of variants and pseudobulk aggregation following recently described best practices [30] (see Methods for details). We applied quasar’s range of models to the OneK1K dataset and, in addition, applied three existing bulk/pseudobulk eQTL mapping methods: the LMM model in apex, the linear model from tensorQTL, and the negative binomial model from jaxQTL [12, 15, 19]. Both the LMM in apex and the negative binomial model in jaxQTL were shown to be the most effective model implemented in their respective packages [15, 19]. To reduce the computational overhead and environmental impact of our comparison we focus our analysis on three cell-types (Plasma cells, “B IN” - immature and naive B cells, and “CD4 NC” - CD4 naive and central memory T cells) that have previously been used as representative cell types for analysis of methods on the OneK1K dataset [14]. The Plasma and CD4 NC are the smallest and largest cell types by total number of cells, respectively, while the B IN cell type is of average size. (Table 1)

**Table 1:** Cell-type sizes in the OneK1K dataset.

| Cell type | Number of cells | Number of individuals |
| --- | --- | --- |
| B IN | 82,068 | 981 |
| B Mem | 48,023 | 981 |
| CD4 ET | 61,786 | 981 |
| CD4 NC | 463,528 | 981 |
| CD4 SOX4 | 4065 | 857 |
| CD8 ET | 205,077 | 981 |
| CD8 NC | 133,482 | 981 |
| CD8 S100B | 34,528 | 980 |
| DC | 8,690 | 968 |
| Mono C | 38,233 | 968 |
| Mono NC | 15,166 | 933 |
| NK | 159,820 | 981 |
| NK R | 9677 | 969 |
| Plasma | 3625 | 795 |

First, we validated quasar by comparing its output to that of existing methods where the statistical model aligns. In particular, we compared quasar’s LM method to tensorQTL, the NB-GLM method to jaxQTL and the LMM method to apex. To assess this concordance we plotted variant level z-scores of quasar methods against z-scores computed by the other methods (Figure 1a). In this analysis, the quasar LM and LMM methods showed very high concordance with tensorQTL and apex respectively, while the NB-GLM showed lower but still good concordance. Overall, the analysis demonstrates that quasar’s output is concordant with that of other widely used eQTL mapping methods.

**Fig 1:**
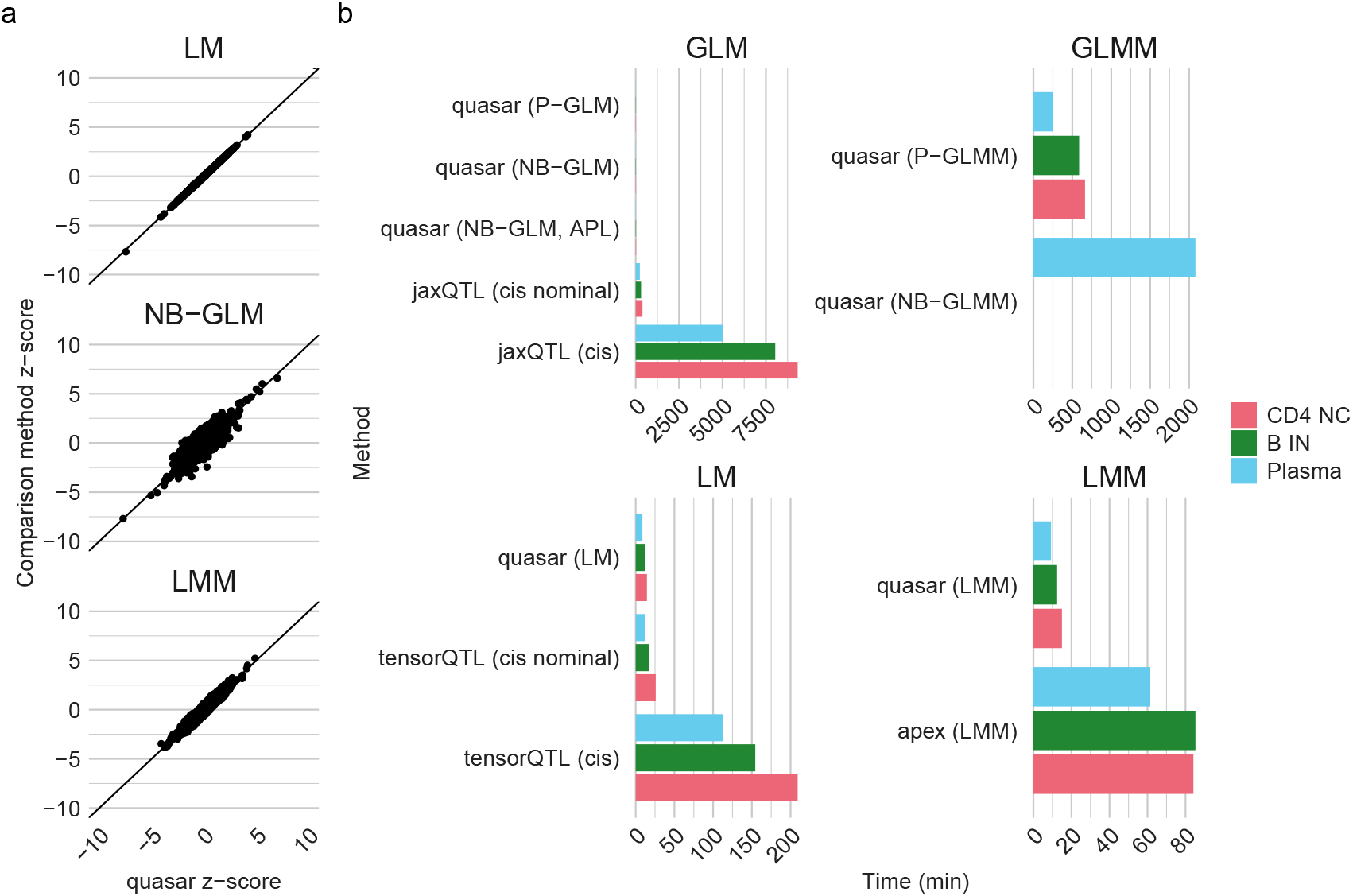
Method concordance and computational performance. a) Scatter plots of variant level z-scores of quasar models against the comparison method that uses the same statistical model (LM: tensorQTL, NB-GLM: jaxQTL, LMM: apex. All results are computed for the B IN cluster, with 100 randomly sampled variants per chromosome. z-scores are restricted to the range (−10, 10) to reduce the impact of outliers on the plots. b) Speed of methods across the three representative cell types. All methods were run on CPUs. Methods are labelled by the options used to run them: for tensorQTL and jaxQTL ‘cis’ computes significance at the level of genes while ‘cis nominal’ computes significance at the level of variants.

While running the negative binomial GLMM, we observed that the estimated values of the dispersion parameter (denoted by *ϕ*) are very close to zero (Figure S2). This observation is consistent with the previous use of heuristic algorithms to better estimate *ϕ* in negative binomial GLMM models and reports that the Poisson GLMM model is sufficient for QTL mapping, as when *ϕ* ≈ 0 the negative binomial GLMM model reduces to the Poisson GLMM model. Theoretically, if the variance-covariance matrix of the random effect is the identity matrix, which would occur if all individuals are completely unrelated, then it can be shown that the Poisson GLMM induces the same form of quadratic mean-variance relationship as a negative binomial model without random effects (see Supplementary Methods for details). This quadratic mean-variance relationship is one of the main factors motivating the use of the negative binomial distribution in modelling RNA-seq data. We also show that in the negative binomial GLMM model, if the random effects are uncorrelated then the dispersion parameter and variance of the random effect are not identifiable, and therefore the standard estimation algorithm will therefore lead to the variance of the random effect accounting for all overdispersion (see Supplementary Methods for details). As individuals in the OneK1K dataset do not show strong relatedness, this effect explains the small *ϕ* values. Therefore, due to its equivalence with the Poisson GLMM when *ϕ* is small we assess the speed of the negative binomial GLMM model but do not otherwise assess it in this paper.

### Computational performance

We next assessed the speed of quasar’s models and compared it to other methods for eQTL mapping. The speed of a method has a large impact on its usability, especially as datasets continue to grow in size. We measured the speed of the methods on the three representative cell types and, for consistency, ran all methods using CPUs, which are generally less expensive to use and more available than GPUs in research and commercial high-performance computing platforms. The quasar package is designed to compute both variant- and gene-level p-values in one run, as is apex, while the tensorQTL and jaxQTL methods compute variant- and gene-level p-values in separate commands. Here, tensorQTL and jaxQTL methods use permutations to compute gene-level p-values while apex and quasar use the ACAT method. Due to its poor computational performance and equivalence with the Poisson GLMM method, we only ran the negative binomial GLMM method on the small Plasma cell type.

First, we compared the computational performance of quasar to the comparison eQTL mapping methods, where the statistical model aligns (Figure 1b). When considering the time taken to compute both variant- and gene-level p-values the quasar models were substantially faster across celltypes when compared to the same model implemented in the comparison eQTL mapping packages: linear model (tensorQTL): 15-19x, negative binomial GLM (jaxQTL): 466-550x, linear mixed model (apex): 5-7x. In addition, quasar was also faster, though to a smaller extent, when comparing its speed to the time taken to compute variant-level p-values only: linear model: 1.4-2x, negative binomial GLM: 22-25x. This finding demonstrates that while a major determinant of quasar’s efficiency is its use of the ACAT test, its variant-level p-value calculation is also more efficient. Second, we compared the computational performance of the different models implemented in quasar (Figure S1)

. The fastest models were the Poisson GLM model and the linear model, followed by the linear mixed model and the negative binomial GLM Other than the GLMM methods, the quasar methods took less than 30 minutes to run on the largest CD4 NC cluster. Using the adjusted profile likelihood in the negative binomial GLM was slower than MLE estimation, at 1.4x slower across cell types. The slowest quasar methods were the GLMM model methods, with the Poisson GLMM being 28-50x slower than the Poisson GLM. This reduced performance is caused by the complexity of the penalised quasi likelihood (PQL) algorithm implemented in quasar for fitting GLMMs (see Methods for details), which involves at each iteration inversion of the relatedness matrix, an *O*(*n*^3^) operation, where *n* is the number of individuals, at each iteration. In the Plasma cluster the negative binomial GLMM was approximately 8 times slower than the Poisson GLMM, demonstrating the increased computational burden of fitting the negative binomial distribution. Finally, as expected all models and methods were generally slower as the number of samples available for the cell type increased, with the mixed model methods showing a substantially larger increase compared to other methods. Overall, these results demonstrate that quasar is more efficient than other eQTL mapping methods, and illustrate the large impact of both statistical model and gene-level significance aggregation strategy on computational performance.

### Type 1 error rate control

We next assessed the ability of methods to control the Type 1 error rate, that is, to not produce false discoveries. To assess this aspect of method performance, we used a permutation-based approach. Using permutations allows us to generate null datasets that maintain properties of the data such as the distribution of counts in the single-cell data and correlation between SNPs, which are challenging to simulate accurately [31]. Specifically, we filtered the data to variants on chromosome 21 (chosen as the chromosome with the fewest variants) and permuted the genotypes, by permuting the individual IDs in the genotype data, 10 times with respect to the gene expression data (see Method for details). All quasar models, except the negative binomial GLMM, and the three comparison methods were then run on the three representative cell types. All quasar and comparison models included sex, age, two expression principal components and six genotype principal components as covariates.

First, we assessed the ability of methods to produce well calibrated p-values when calculating p-values for variants (Figure 2). Across quasar models, the LM and LMM displayed a uniform distribution of p-values, and were slightly conservative for very small p-values. The two quasar NB-GLM models showed overall good calibration with slightly-too-small p-values when the p-values were small in the small Plasma cluster, though this was reduced when using the adjusted profile likelihood method for dispersion estimation. While performing this analysis we noticed that genes with a very large variance in counts relative to the mean caused false positives for the negative binomial methods. Therefore, in this and other analyses, we filtered out the top 0.5% of genes ranked by coefficient of variation for all count-based methods (Figure S4).The Poisson GLMM model displayed uniform p-values for the B IN and CD4 NC cell types but had a small excess of small p-values for the small Plasma cluster. Across all cell types the Poisson GLM produced very poorly calibrated p-values, demonstrating that it could not robustly control Type 1 error. This observation is consistent with previous work and is likely caused by the Poisson distribution’s inability to model overdispersion, unlike the negative binomial model [15]. Due to its failure to control the Type 1 error rate we do not use the model in other comparisons and running quasar specifying the Poisson GLM generates warnings.

**Fig 2:**
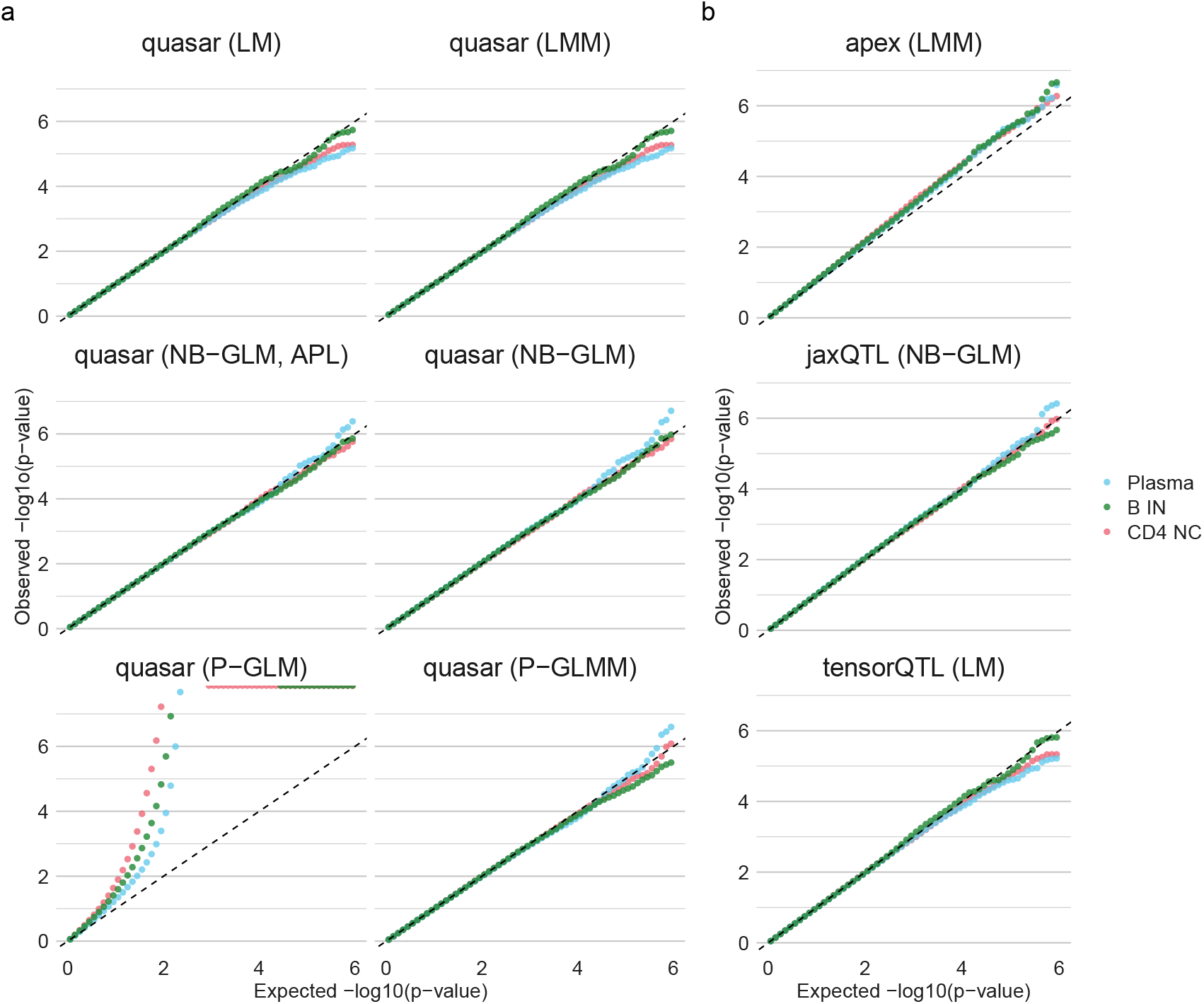
Distribution of variant-level p-values under a permutation null. a) Results across the statistical models implemented in quasar, showing results for the three representative cell types. b) Results for the comparison eQTL mapping methods, also across the three representative cell-types. N = 75,395 SNPs tested, binning was performed for visualisation purposes.

Considering the three comparison methods, tensorQTL produced uniform p-values across all cell types while jaxQTL showed results similar to the the quasar NB-GLM model. In our hands, apex produced overly small p-values across all cell-types, possibly due to issues with processing of the genetic relatedness matrix (see Methods for details). Next, to more formally assess the Type 1 error control of the methods, we computed the proportion of variants declared significant at different significance thresholds (Figure S5a). This analysis highlights the ability of the adjusted profile likelihood dispersion estimation to improve the calibration of the p-values. Even at large *α* thresholds the NB-GLM methods (in quasar and jaxQTL) did not produce calibrated p-values, but the NB-GLM estimation with the adjusted profile likelihood was well calibrated. This improved calibration is due to the less biased estimation of the dispersion when using the adjusted profile likelihood, leading to generally larger values of the dispersion, increasing the estimated variance (Figure S3).

Second, we assessed the ability of methods to produce well calibrated p-values at the level of genes (Figure 3). All quasar methods use the ACAT method to compute per-gene p-values. The LM and LMM quasar methods produced calibrated results, similar to the tensorQTL results attained using permutations. The quasar NB-GLM methods displayed had p-values that were slightly too small, especially when the computed p-values were small but the use of the APL greatly improve calibration. For the comparison methods, jaxQTL and tensorQTL showed well calibrated distributions while apex had an excess of small p-values. The poor performance of apex at the gene-level shows that the ACAT method’s performance is highly dependent on the calibration of variant-level p-values. In addition, we computed the proportion of variants declared significant at different significance thresholds (Figure S5b). This analysis highlighted similar results to the p-value calibration analysis but while the adjusted profile likelihood improved performance, the negative binomial GLM still displayed a slightly inflated Type 1 error rate for the stricter significant threshold (*α* = 0.01).

**Fig 3:**
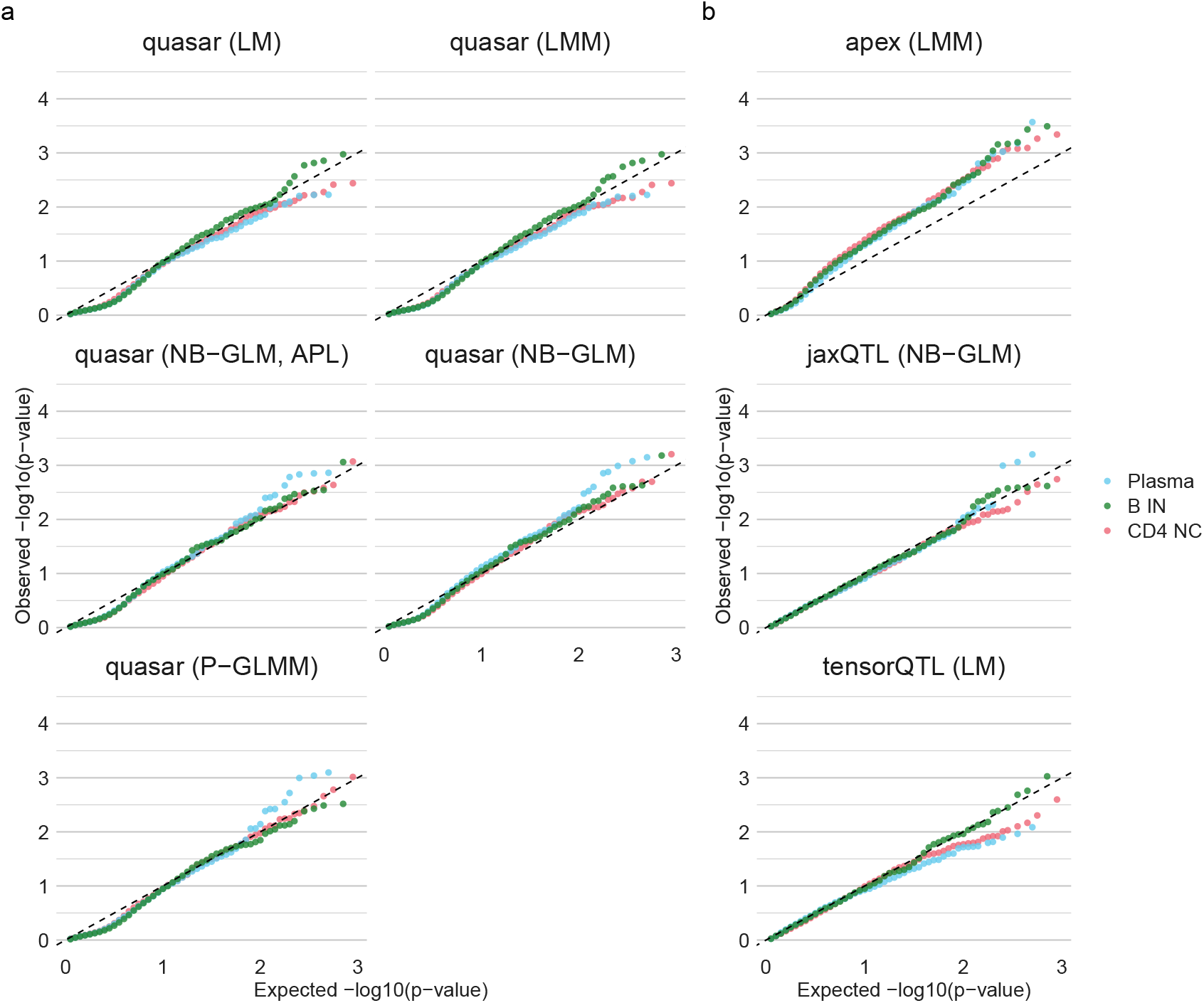
Distribution of gene-level p-values under a permutation null. a) Results across the statistical models implements in quasar, showing results for the three representative cell types. b) Results for the comparison eQTL mapping methods. N = 75,395 SNPs tested, aggregated into gene-level p-values. Binning was performed for visualisation purposes.

While performing these analyses, we noticed that the distribution of ACAT p-values under the permutation null was substantially non-uniform, instead displaying a bi-model distribution (Figure S6). This distribution is different to that of permutation-computed p-values which do show the expected uniform distribution (Figure S6b). We confirmed that this pattern is not due to issues in quasar, as it occurs when ACAT is computed from the output of tensorQTL (as in the figure) and it can also be observed in log-transformed Q-Q plots in other methods that use the ACAT, as a small bump below the line when the p-values are close to one. The bimodal distribution under the null does not satisfy the assumptions of the q-value estimation procedure, which has previously been used to assess the significance of gene-level p-values computed using permutations [32]. We further investigated the effect of this misspecification when comparing the power of methods. Overall, however, we found that the ACAT method displayed uniform p-values under a permutation null in the left tail, demonstrating that the ACAT distribution is well-calibrated under the null for the small p-values of interest in eQTL mapping analysis.

### Power analysis

Finally, we assessed the statistical power of all methods. We assessed power at the level of both variants (how many eQTLs a method identifies) and genes (how many eGenes a method identifies). To quantify the the power of methods to detect eQTLs, we computed the number of variants that are significant at a *p <* 5 *×* 10^−6^ threshold (Figure 4a). In the three representative cell types, we observed that count-distribution methods (quasar NB-GLM with and without the APL, Poisson GLMM, and jaxQTL) displayed the greatest power to detect eQTLs, as previously described [15]. In the Plasma cell type the NB-GLM methods of quasar and jaxQTL method mapped more eQTLs than the quasar NB-GLM (APL) and P-GLMM methods, highlighting the improved calibration of the latter two approaches. The next most powerful type of methods were linear model methods, with LMM-based methods surprisingly showing the lowest power. For the the LM and LMM models implemented in quasar, the number of significant variants was highly concordant with the tensorQTL and apex methods, respectively.

**Fig 4:**
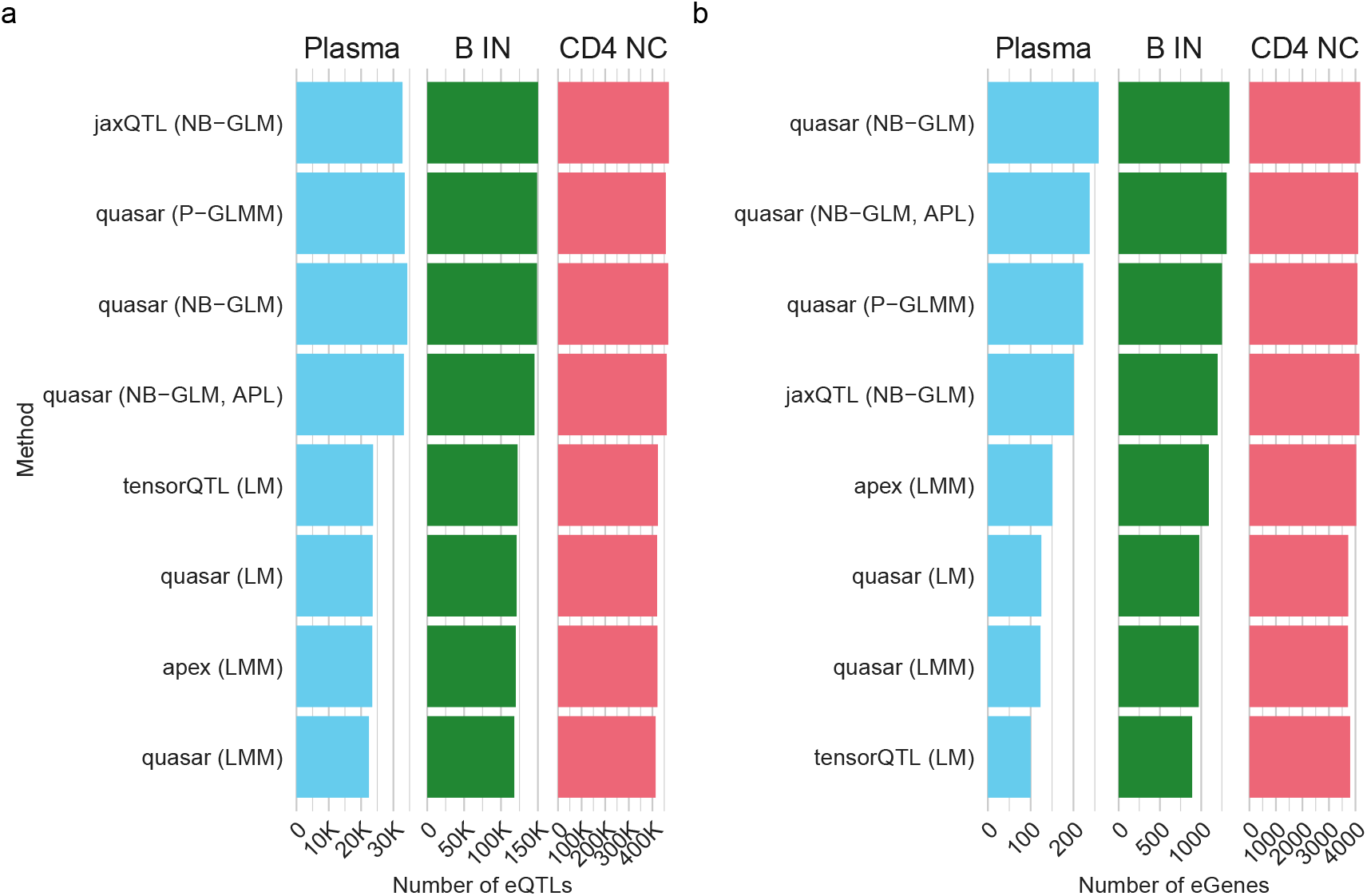
Statistical power of all methods in the three representative cell types. a) The number of eQTLs, at a *p <* 5 *×* 10^−6^ threshold, found across the three representative cell-types by quasar, with different models, and the comparison eQTL mapping methods. b) The number of eGenes, at a 5% FDR threshold, found across the three representative cell-types by quasar, with different models, and the comparison eQTL mapping methods.

Next, we quantified the power of methods to detect eGenes, computing the number of genes which were significant at a 5% FDR threshold (Figure 4b). The power of methods at the gene level is affected by both the model and gene-level aggregation procedure and the ACAT test has been previously been shown to have greater power than the permutation approach [19]. To assess the impact of ACAT in our data and study the use of q-values with ACAT, where assumptions of its estimation procedure are not met, we applied all combinations of procedures to the output of tensorQTL (Figure S7a). This analysis had three main findings. First, as previously reported, the ACAT method generally detected more eGenes than the permutation-computed p-values [19]. Second, applying the Benjamini-Hochberg (BH) method and the q-value procedure to the ACAT p-values produced the same set of eGenes. Third, when the BH and q-value procedure were applied to the permutation p-values, the q-value procedure detected more eGenes and the increase was larger for the larger cell types. Reassuringly, the overall overlap between all sets was very high (Figure S7b). Based on these findings, we recommend that the BH procedure be used with ACAT p-values, as the technical assumptions of the q-value estimation procedure are not met and the methods produce equivalent results. However, for the permutation approaches our results validate the use of the q-value method as increasing power. Therefore, in our overall comparison of methods we use the BH procedure for methods that compute ACAT p-values and the q-values procedure for methods that compute permutation p-values.

Overall, the gene-level results were concordant with the variant-level results with count-distribution methods again displaying higher power. This relative increase in power decreased as the sample size of the cell type increased. The apex method displayed a much higher power than in the variant-level analysis but this effect is due to its poor calibration under the null, highlighting the ACAT method’s tendency to produce false discoveries if the variant-level p-values are not well calibrated. As the sample size of the cell type increased the methods using q-value corrected permutation p-values— jaxQTL and tensorQTL—had higher power. Indeed, while they had lower power in the Plasma and B IN cell types, in the CD4 NC cell type (the largest in the OneK1K dataset) both methods had comparable or slightly higher power than the equivalent quasar method.

To confirm that the quasar models were finding a concordant set of eGenes, we visualised the overlap between the eGenes selected by different models on the B IN cell type (Figure S8a). This analysis showed that, as expected, most eGenes were found by all models with a subset being found only by count-distribution methods. The eGenes only identified by the count-distribution methods had lower total counts compared to eGenes identified by all methods (*p <* 5.2 *×* 10^−4^, two sided Wilcoxon rank sum test; Figure S8b). This observation, which is in line with previous literature, suggests that the count-distribution methods’ higher power can be attributed to better performance in genes with lower expression.

Finally, we sought to expand our analysis of the finding that count-distribution methods have more power for eQTL mapping. To enhance our analysis we additionally ran the quasar linear model and quasar negative binomial GLM with the adjusted profile likelihood in the 11 other cell types in the OneK1K dataset (Figure 5). As in the previous analysis we computed the number of eQTLs and eGenes found by each method. The negative binominal GLM had more power than the linear model across all cell types. Overall, across cell types the negative binomial GLM found 23% more eQTLs and 29% more eGenes. The two monocyte cell types showed the largest gain in power for the negative binomial GLM method.

**Fig 5:**
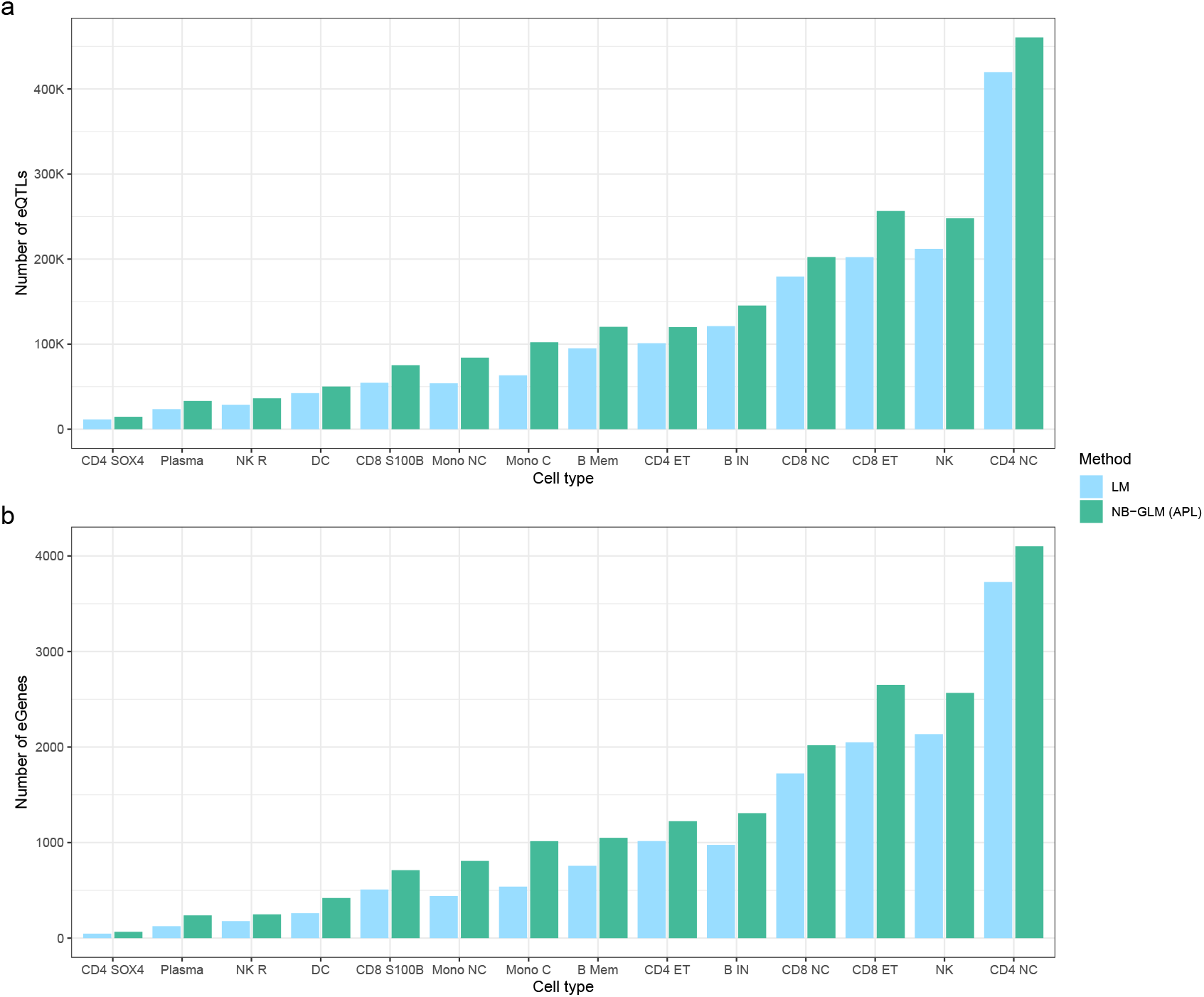
Statistical power of negative binomial and linear model methods across the OneK1K dataset. a) Number of eQTLs detected by the quasar linear model and negative binomial GLM with adjusted profile likelihood dispersion estimation methods across all cell types in the OneK1K dataset. b) Number of eGenes detected by the quasar linear model and negative binomial GLM with adjusted profile likelihood dispersion estimation methods across all cell types in the OneK1K dataset.

## Discussion

In this paper we introduced quasar, a flexible and efficient C++ command line program for performing eQTL mapping. Compared to existing eQTL mapping methods, quasar implements a wider variety of statistical models, including both count distribution and mixed models. Here, we also introduce and implement in quasar an analytic approximation to the score-test variance in mixed models. Furthermore, we highlighted that difficulties in estimating the negative binomial dispersion, identified in the context of RNA-seq differential expression testing, also apply to eQTL mapping. Therefore, quasar implements the Cox-Reid adjusted profile likelihood, which enables unbiased estimation of the dispersion. We demonstrated quasar in a pseudobulk analysis of the OneK1K dataset and compared it to three existing eQTL mapping tools: tensorQTL, jaxQTL and apex.

The quasar software package has several advantages relative to existing eQTL mapping software. Compared to tensorQTL, quasar’s linear model implementation gives highly similar results and is up to 2x faster for computing variant-level p-values on CPUs and replaces permutations with the ACAT method. Compared to jaxQTL, quasar’s negative binomial GLM gives highly similar results, is around 40x faster on CPUs for computing variant-level p-values and additionally implements the adjusted profile likelihood for estimating the negative binomial dispersion. Finally, compared to apex’s linear mixed model, quasar’s linear mixed model implementation is faster and provides robust control of Type 1 error.

We exploited quasar’s flexibility to compare different statistical models for eQTL mapping without confounding due to other method implementation details. In this analysis, and consistent with previous reports, we observed that count-distribution approaches have more power, with around 30% more eGenes found across cell-types. We observed that the negative binomial GLM could fail to produce well-calibrated p-values and control the Type 1 error rate at the variant level but that this effect could be fixed by using the adjusted profile likelihood to estimate the dispersion parameter. Furthermore, in the datasets we considered, mixed models did not display better performance but were substantially slower. Overall, we recommend the use of the negative binomial model with the adjusted profile likelihood. The linear model can also be used as it is slightly faster than the NB-GLM at the cost of reduced power. Mixed model approaches should be considered if there is known to be substantial relatedness between samples. In this scenario, the speed of quasar makes conducting a permutation-null study of different models Type 1 error characteristics highly feasible, enabling dataset-specific assesment of model performance.

Our conclusions about the utility of mixed models are specific to current datasets. It is possible that in larger datasets, where cryptic relatedness is more likely, or in datasets that are known to contain related individuals, mixed models could have better performance. We encourage users of quasar to exploit its flexibility and try mixed models in these situations. However, our conclusions should hold in all datasets with low relatedness and less than 1000 individuals, which accounts for a large proportion of all current eQTL datasets. We also note that analysis of Phase 1 of the TenK10K project, including approximately 2000 individuals, did not use the ability of SAIGE-QTL to model sample relatedness [8]. Mixed models became standard in GWAS analysis as sample sizes increased to biobank scale so the performance of mixed models will likely need to be revisited as larger eQTL datasets, and datasets collected from established biobanks, become available. Furthermore, our findings about the use of mixed models for relatedness are not relevant to the use of mixed models to model the repeated measurement structure of single cells nested within individuals.

Our findings have implications for the broader landscape of QTL mapping methods. First, we observed empirically and derived theoretically that in situations with low relatedness Poisson GLMMs can model a quadratic mean-variance relationship. This finding justifies the lack of negative binomial model in SAIGE-QTL and suggests negative binomial mixed models may be broadly superfluous except in situations with known strong sample relatedness. Second, our use of ACAT across all methods in quasar highlights its wide applicability and we showed that using ACAT increased power. This increase may reflect ACAT’s use of multiple SNPs, while the permutation approach considers only the minimum p-value per gene. Overall, we recommend that ACAT can be widely applied in place of permutations in eQTL mapping. However, we note that ACAT does not have a uniform distribution under the null, breaking the assumptions of the q-value fitting procedure, so we instead recommend Benjamini-Hochberg FDR correction be used to perform multiple testing correction when the ACAT is used. Third, while in this paper we focused on eQTL mapping, quasar is likely to be effective for mapping other molecular QTLs. For example, chromatin accessibility QTLs could be modelled using the negative binomial model as ATAC-seq data is known to follow a count distribution. Furthermore, quasar could be used to map other types of QTLs such as cell-state or cell-state abundance QTLs [33, 34].

Finally, quasar has some limitations. First, quasar is not designed for larger, biobank-scale datasets (approximately *>* 20, 000 individuals), where direct inversion of the genetic relatedness matrix required when fitting the mixed models is not computationally feasible. To scale to such datasets quasar could implement the preconditioned gradient descent algorithm used for matrix inversion by SAIGE and SAIGE-QTL. Second, unlike apex and SAIGE-QTL, quasar does not support sparse or low-rank GRMs. Third, unlike jaxQTL and tensorQTL, quasar does not currently have the ability to accelerate performance with GPUs if they are available. Integration of OpenCL would allow quasar to also use GPUs to accelerate its performance. Finally, quasar is designed to work on bulk or pseudobulk single-cell data, unlike SAIGE-QTL, which models single-cell level counts directly. Extending quasar to efficiently map eQTLs at single-cell resolution is a promising direction for future research. A limitation of our comparison of methods was that we were unable to run jaxQTL and tensorQTL with GPUs due to computational constraints.

Overall, quasar provides a highly performant and flexible command line software tool for performing bulk and pseudobulk eQTL mapping.

## Methods

### Overview of quasar

The quasar method uses the following overall algorithm. (More details about the overall algorithm and all aspect of the quasar’s methodology are available in the Supplementary Methods)

1. For each gene, quasar fits a ‘null’ model where the measured counts or transformed gene expression values are modelled as a function of nuisance covariates – such as age, sex or genotype PCs – supplied by the user.
2. Then, for each gene, the estimated model parameters are used to remove the effect of the nuisance covariates on the gene expression values.
3. Next, all genetic variants tested for association have the effect of the the nuisance covariates removed. That is, we regress the genotype vector on the nuisance covariates and subtract the predicted values.
4. Finally, for each genetic variant-feature combination, a statistical test measuring association (specifically, a score test) is performed using the processed features and genetic variants described above.

### Statistical models

The quasar software implements the following statistical models for residualising the phenotype:

- Linear model,
- Poisson generalised linear model,
- Negative binomial generalised linear model,
- Linear mixed model,
- Poisson generalised linear mixed model and
- Negative Binomial generalised linear mixed model.

To fit the linear model we use the standard analytic MLE. To fit the Poisson GLM we use the standard iteratively reweighted least squares (IRLS) algorithm, the same algorithm used by the R function `stats::glm()`. To fit the NB GLM we use the standard algorithm, as implemented in the R function `MASS::glm.nb()`, that alternates between estimating the dispersion parameter *ϕ* and fitting a NB GLM where *ϕ* is known. To fit the LMM we use the FaST-LMM algorithm [35], with the C++ implementation based on that of apex. To fit the Poisson GLMM we use the penalized quasi-likelihood algorithm (PQL), which has been previously applied to fit logistic GLMM for GWAS SAIGE and for single-cell level QTL mapping in SAIGE-QTL. The algorithm has also been applied to estimate heritability for count-distributed responses with PQLSeq/PQLSeq2 and we use methodological elements of the PQLSeq algorithm in quasar [36] (See Supplementary methods for details). To fit the NB GLMM we use an analogous algorithm to that used for the NB GLM, replacing the Poisson GLM and Negative Binominal GLM with *ϕ* fixed with their mixed model equivalents. This is the same algorithm used in the R function glmer.nb(), from the widely used lme4 package for mixed models, which has previously been used to map single-cell eQTL [5, 37]. All counts models in quasar include an offset term is computed are the total counts for each individual across genes.

### Score test variance approximation

When mixed models are used to residualise the feature outcome the variance of the score statistic has the form

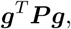

where ***g*** is the *n*-vector of genotypes after covariate residualisation and ***P*** is a dense *n × n* residualisation matrix that arises in the mixed model fitting procedure (See the Supplementary Methods for details). This quantity must be computed for each variant, but this is not feasible as computing it has computational complexity *O*(*n*^2^) and *n*, the number of variants, may be large. This computational problem has been addressed by approximating the score test variance with approximations of the form

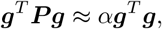

where the constant *α* is chosen to make the approximation as accurate as possible. (Here we consider the approximation in the LMM case; for the GLMM case see the Supplementary Methods.) The most common approach for choosing *α* is the variance ratio approach, also known as the GRAMMAR-GAMMA approximation where we set

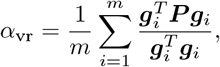

where *m* is generally taken to be small (≈ 30) and the variants, ***g***_*i*_, are drawn at random genome-wide [27]. This approach is now used across GWAS and eQTL methods [14, 21]. A different approach, used by the software package for GWAS analysis regenie, is to set *α* as

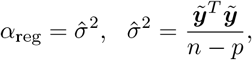

where 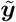 is the value of the response after nuisance covariates have been regressed out, *n* is the number of individuals and *p* is the number of nuisance covariates [28]. The regenie estimator is also closely related to the original GRAMMAR estimator [38]. Here, we propose to set *α* using the trace of ***P***; specifically, we propose

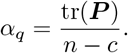

This expression is an approximate expectation of the variance-ratio approximation, treating ***g*** as random. (See the Supplementary Methods for a derivation.) This new estimator can be calculated from ***P*** in *O*(*n*), removing the need to perform any *O*(*n*^2^) complexity ***P g*** multiplications. In the case of the GLMM, the need for a second-order correction means the trace-based approximation also has computational complexity *O*(*n*^2^); see the Supplementary Methods for details.

This novel estimator also helps to clarify why *α*_reg_ ≈ *α*_vr_, despite appearing to estimate different quantities. For a LMM, we show that under null of the genetic variant having no effect on the feature

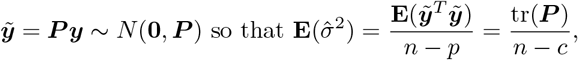

so the regenie approximation is effective because it estimates the trace of ***P***. In quasar, we use the regenie approximation for the LMM case as the full ***P*** matrix is not explicitly formed by the algorithm used to fit the LMM. For the GLMM methods we use the novel approximation (described in detail in the Supplementary Methods.)

### Adjusted profile likelihood

In a frequentist setting the negative binominal dispersion parameter, *ϕ*, is estimated using maximum likelihood. However, the maximum likelihood estimator of *ϕ* is biased, systematically underestimating the dispersion [16]. This issue has been previously identified in the context of differential expression testing in RNA-seq data, where instead maximising the Cox-Reid adjusted profile likelihood has been shown to alleviate it [17, 25]. The quasar package implements the option to estimate the dispersion parameter *ϕ* in the NB GLM and NB GLMM models by maximising the adjusted profile likelihood, which is the penalized log-likelihood,

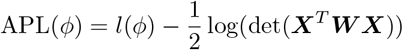

where *l*(*ϕ*) is the usual log likelihood, det denotes the determinant, and ***X***^*T*^ ***W X*** is the information matrix, where ***X*** is the matrix of covariates and ***W*** is the diagonal matrix of working weights. We maximise APL(*ϕ*) using Brent’s algorithm, which is chosen for convenience as it is also used to fit the LMM.

### Aggregated Cauchy association test

To compute gene-level p-values quasar uses the aggregated Cauchy association test (ACAT) method [24]. The ACAT method is a meta-analysis approach specifically designed to handle situations where the p-values are highly correlated, as in eQTL mapping. The ACAT was first used for this purpose in APEX and is also used by SAIGE-QTL [14, 19]. Prior to the use of the ACAT, a permutation approach was used [12], however this is computationally expensive. For a vector of p-values *p*_1_, …, *p*_*k*_ the ACAT statistic is defined as

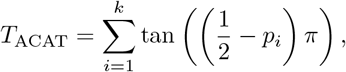

where tan(·) is the standard tan function. If *p*_*i*_ ~ *U* (0, 1), i.e., under *H*_0_, then 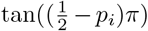 follows a standard Cauchy distribution. Under *H*_0_, the distribution of *T*_ACAT_ is well approximated by a Cauchy distribution with location parameter 0 and scale parameter *k*. Therefore, the associated p-value of the *T*_ACAT_ statistic, *p*_ACAT_, is approximately

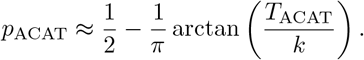

### OneK1K dataset processing

The variant data was processed in accordance with previous descriptions [6]. We filtered the variant data to only variants with MAF *>* 0.05 and passing a Hardy-Weinberg equilibrium test at *p >* 10^−6^. Following recommendations for pseduobulk eQTL analysis [30], for methods that require count data as input we compute the sum for each individual of the raw counts, while for methods that require normalised input we normalise the counts with Scanpy and then compute the mean for each individual. We performed this process for each cell type, in each cell type filtering to genes where at least 10% of counts were non-zero. This filtering step only requires that 10% of counts are non-zero in the cell-type of interest, not in the whole dataset. For the genotype data, we filtered to variants with INFO *>* 0.8, MAF *>* 0.05 and Hardy-Weinberg equilibrium p-value *>* 1 *×* 10^−6^. To get genelevel information for analyses we used the gene annotation GTF file (Homo sapiens.GRCh37.82) and collapsed it to a single transcript model using “collapse_annotation.py” from GTEx analysis pipeline. In addition, we excluded a small subset of genes with very large variance in the observed counts, which caused spurious false positives in the negative binomial models (both jaxQTL and quasar). Specifically, we exclude the top 0.5% of genes ranked by coefficient of variation computed from the raw counts from all count-based analysis. The coefficient of variation is defined as the standard error divided by the mean.

### Computation of genetic relatedness matrices

To compute the genetic relatedness matrix for use in quasar mixed model methods we filtered the data to remove variants in LD using the command `plink2 --indep-pairwise 250 50 0.2` then computed the GRM using the recommended `plink2 --make-king` command. To ensure the computed GRM was positive definite we used the R function `Matrix::nearPD()` to compute the nearest positive definite matrix to the matrix returned by plink and used that matrix as the GRM. We computed one GRM across all chromosomes. Initially, we used the same GRM in apex, however with this estimated GRM apex produced a very large number of false positive variants with small p-values. While trying to fix this issue we observed that apex performed better when we instead used the GRM estimated with `plink --make-rel`, so we used this GRM with apex. We note that the `--make-king` is recommended over `--make-rel` in the plink documentation as it is more robust to population stratification. As the code used to run apex on real data was not released with the paper describing apex we were unable to determine with which GRM apex’s authors intended it to be used.

### Covariates for QTL mapping

We used sex, age, 2 expression principal components and 6 genotype principal components as covariates for eQTL mapping, following the covariates used in the original OneK1K paper. The genotype PCs were computed using plink after filtering to variants using the command `plink2` `--indep-pairwise250 100 0.3`. The expression PCs were computed using scanpy applied to the pseudobulked Scanpy-normalised gene expression data. Our use of principal components to correct the expression data follows recent recommendations for pseduobulk eQTL mapping [30].

### Comparison QTL mapping methods

#### tensorQTL

We ran tensorQTL in both `cis` and `cis_nominal` modes with all parameters at their default values. The pseudobulked gene expression data was quantile-normalised to standard a normal distribution before being input to tensorQTL.

#### jaxQTL

We ran jaxQTL using the nb model, which was found by the authors of jaxQTL to have the highest power. Following the package documentation, we ran parralelised jaxQTL by running it on chunks of 100 genes. Following the example code, we used 1000 permutations to define gene-level p-values. We changed the default window size to 1000000 (1Mb), as in apex, tensorQTL and quasar.

#### apex

We ran apex using the LMM mode, passing a GRM with the `--grm` flag and also running with the `--rankNormal` flag to quantile normalise the transformed expression values to a standard normal distribution. The apex program only implements the ACAT method to compute gene-level p-values. As described above, we used a GRM computed with `plink --make-rel`.

### Permutation analysis

For the permutation analysis we subsetted the data to chromosome 21 and permuted the data 10 times. To permute the plink data we permuted the ids in the `.fam` file using R, while for the vcf we permuted the sample IDs using `bcftools` and the `shuf` command. While most methods take plink.bim/.bed/.fam as input the apex methods required.vcf file as input. This analysis was performed for each of the three representative cell types. We visualise the results of the permutation analysis using a quantile-quantile plot of the −log10 transformed p-values.

### Power analysis

To quantify the power of the methods we computed:

- The number of eQTLs, defined as variants with p-values *<* 5 *×* 10^−6^;
- The number of eGenes, defined as genes with p-values with Benjamini-Hochberg FDR *<* 0.05 for methods with ACAT p-values and genes with q-values *<* 0.05 for methods with permutation p-values.

We used the `p.adjust()` function with method = “BH” to compute the Benjamini-Hochberg FDR values and the `qvalue()` function to compute q-values.

## Data availability

Single-cell RNA-seq and genotype data of OneK1K are available via Gene Expression Omnibus (GSE196830). Single-cell gene expression data we used the RNA-seq data available on Human Cell Atlas: https://cellxgene.cziscience.com/collections/dde06e0f-ab3b-46be-96a2-a8082383c4a1. Processed genotype data were provided by the authors of the OneK1K paper.

## Code availability

The quasar package is available at: https://github.com/jeffreypullin/quasar. Documentation and instructions for installing and running quasar are available at: https://jeffreypullin.github.io/quasar/. Code used to reproduce the analysis described in this paper is available at: https://github.com/jeffreypullin/quasar-paper.

We downloaded jaxQTL from GitHub https://github.com/mancusolab/jaxqtl/, commit f4f52a. We downloaded tensorQTL from Github at https://github.com/broadinstitute/tensorqtl, commit 812040. We downloaded apex from Githubat https://github.com/corbinq/apex, commit 63b605.

## Acknowledgements

We thank members of the Wallace group for helpful discussions, particularly Andy Bass for discussion about the assumptions of *q*-value estimation procedures. We thank Joseph Powell for his assistance with the OneK1K dataset.

## Supplementary Figures

**Supplementary Figure S1:**
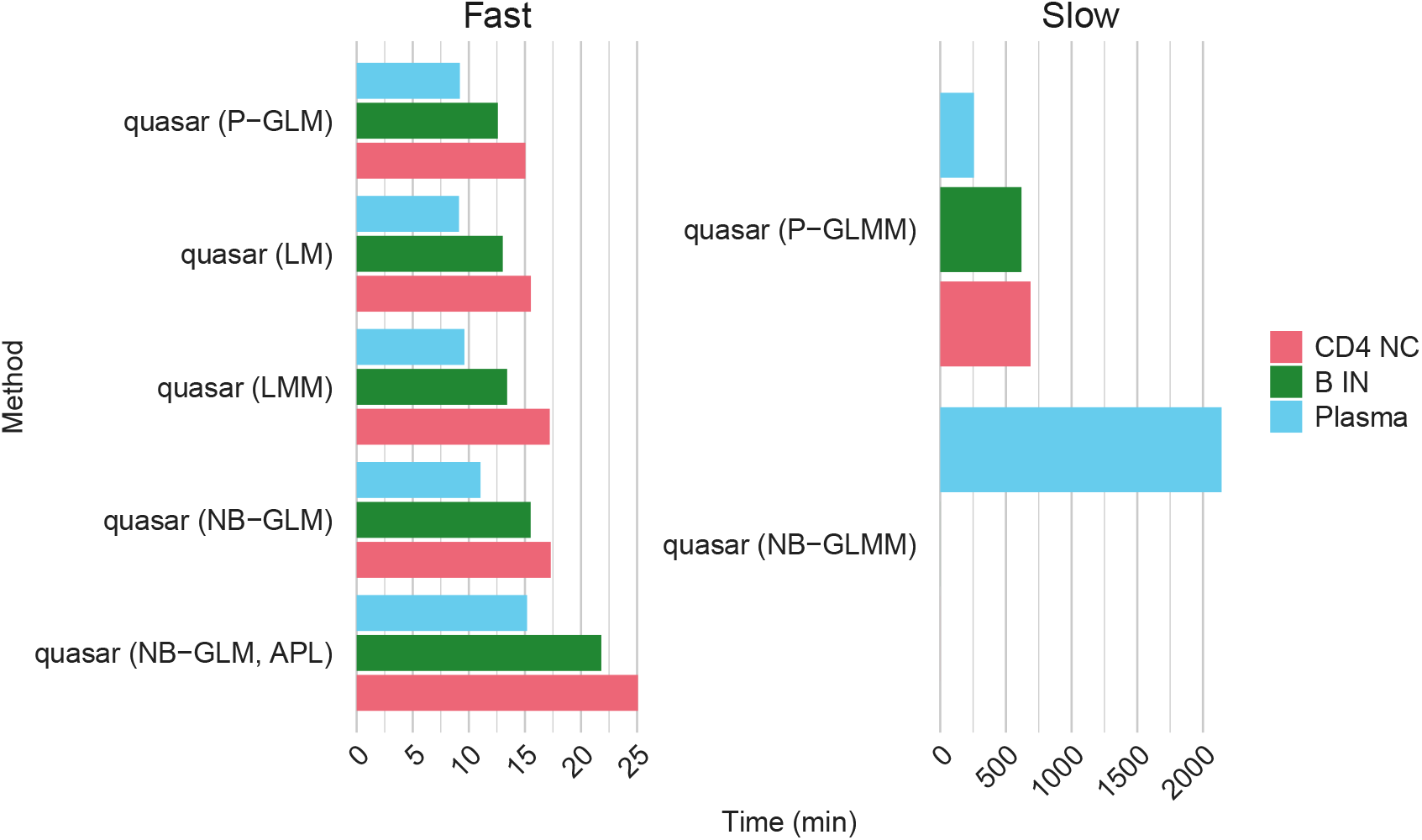
Time to run all quasar methods on the three representative cell types. Time in minutes to run each method across all chromosomes on the three representative cell-types. The methods are faceted by their overall speed for clarity. The NB-GLMM method was not run on the B IN or CD4 NC celltypes.

**Supplementary Figure S2:**
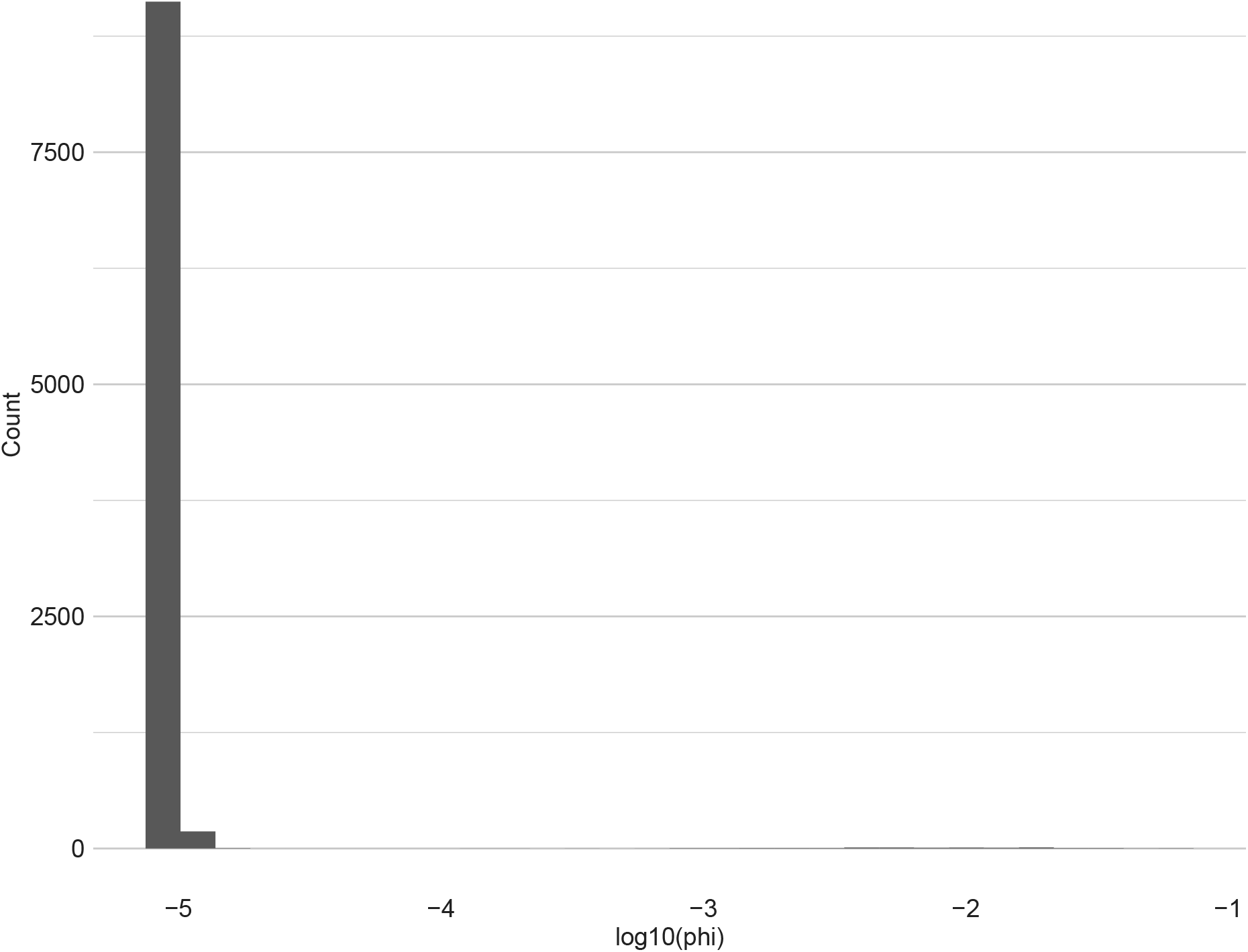
Distribution of the negative binomial dispersion estimates in the NB-GLMM model. Distribution of the estimated negative binomial dispersion, *ϕ*, for the negative binomial GLMM in the Plasma cell type. Almost all values are very small.

**Supplementary Figure S3:**
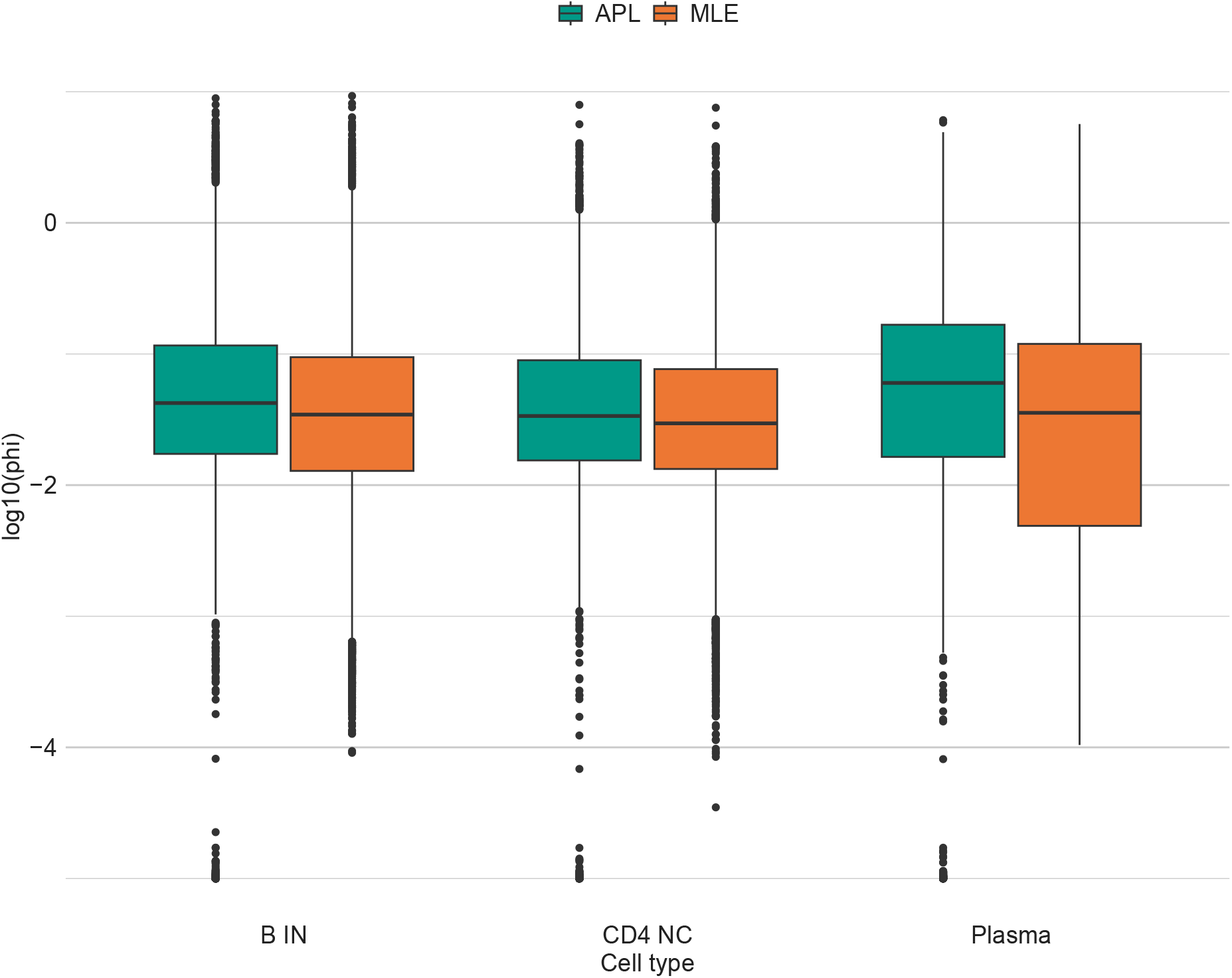
Distribution of negative binomial dispersion estimates with estimation methods. log_10_ transformed values of the negative binomial dispersion parameter, *ϕ*, for genes across the three representative cell types, estimated using both the Cox-Reid adjusted profile likelihood and maximum likelihood.

**Supplementary Figure S4:**
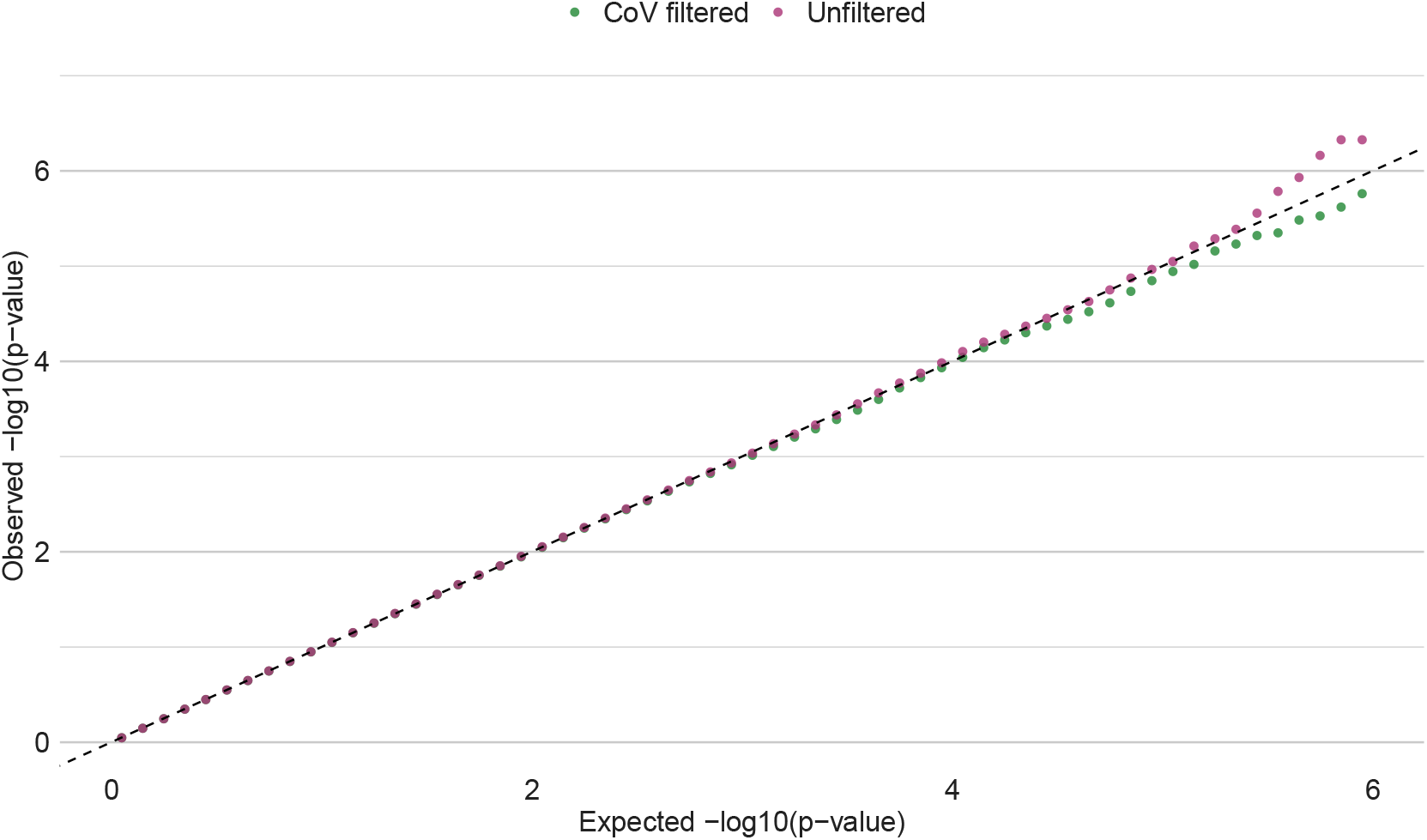
Distribution of variant-level p-values under a permutation null with and without coefficient of variation filtering for the CD4 NC cell-type. As the p-values get small, the unfiltered data displays an inflation of small p-values relative to the expectation, while the filtered data does not. N = 75,395 SNPs tested, binning was performed for visualisation purposes.

**Supplementary Figure S5:**
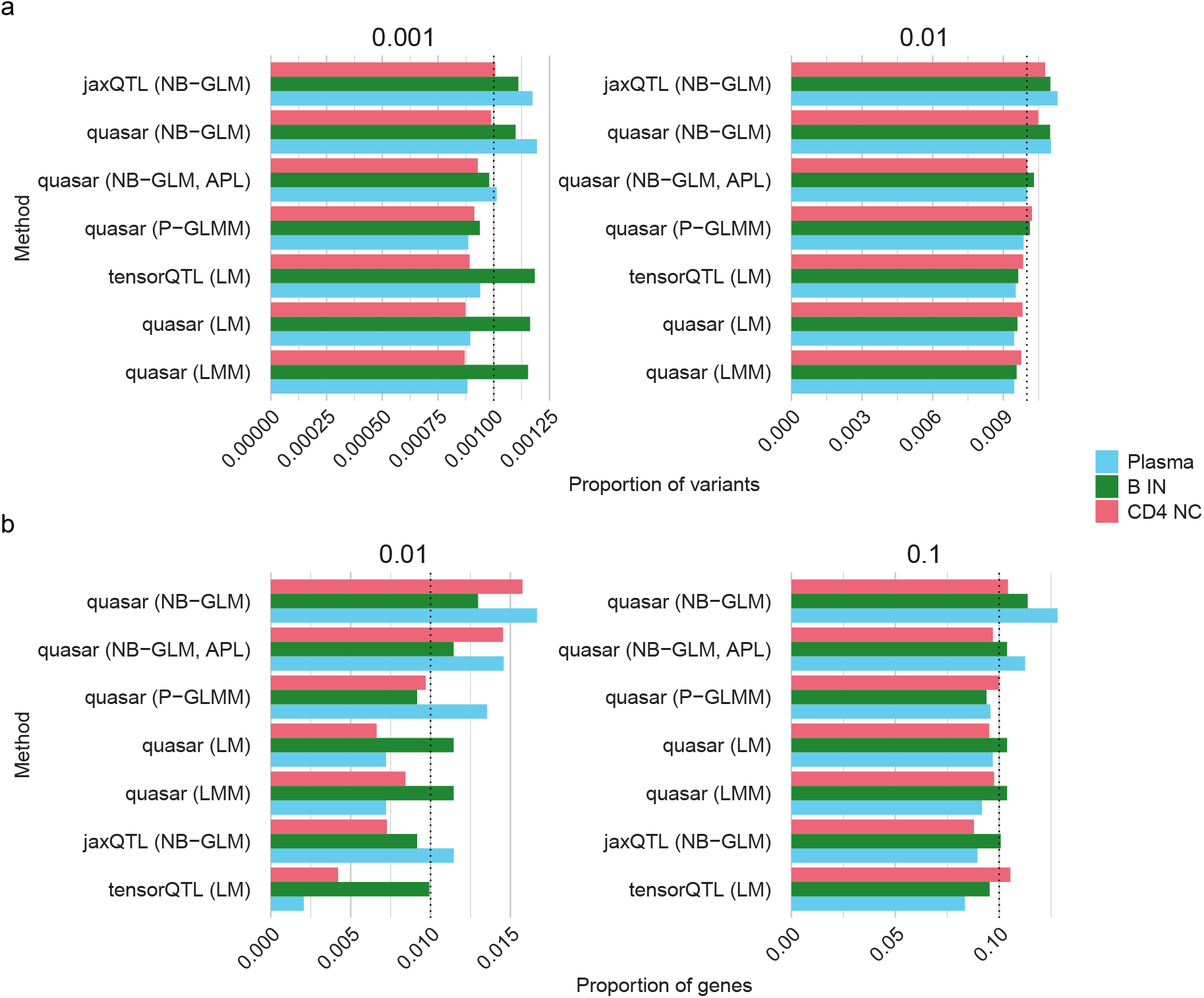
Empirical significance proportions at different thresholds. a) Proportion of significant variants at *α* = 10^−3^, 10^−2^ under a permutation null for the three representative cell types. b) Proportion of signifiant genes at *α* = 10_−2_, 10^−1^ under a permutation null across the three representative cell types. The dotted lines show the *α* level.

**Supplementary Figure S6:**
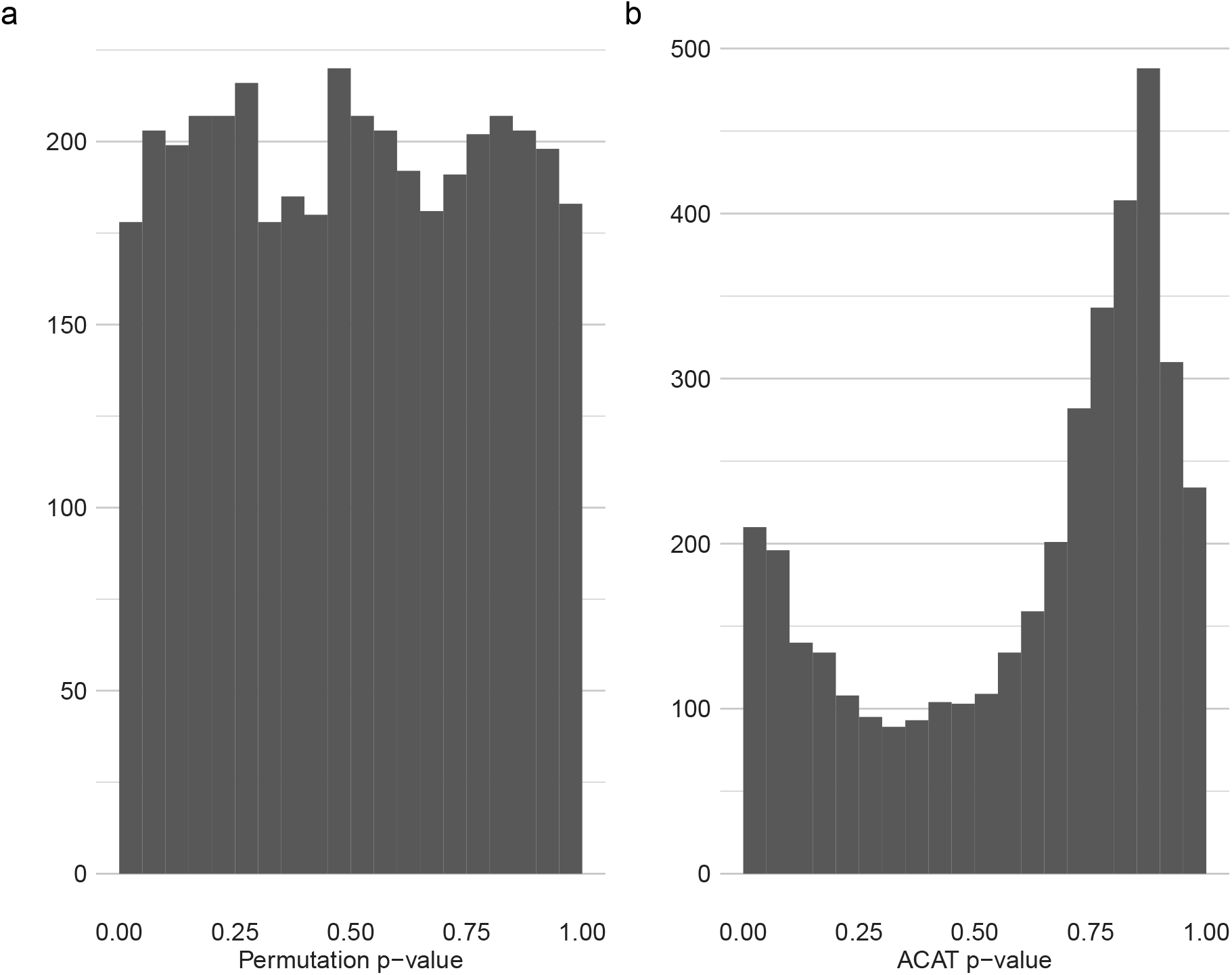
Distribution of gene-level p-values under a permutation null. a) p-values calculated by deriving a permutation p-values for the minimum per-gene p-value b) p-values calculated using ACAT applied to on variant-level p-values. Both panels show the results for tensorQTL run on all three representative cell types combined.

**Supplementary Figure S7:**
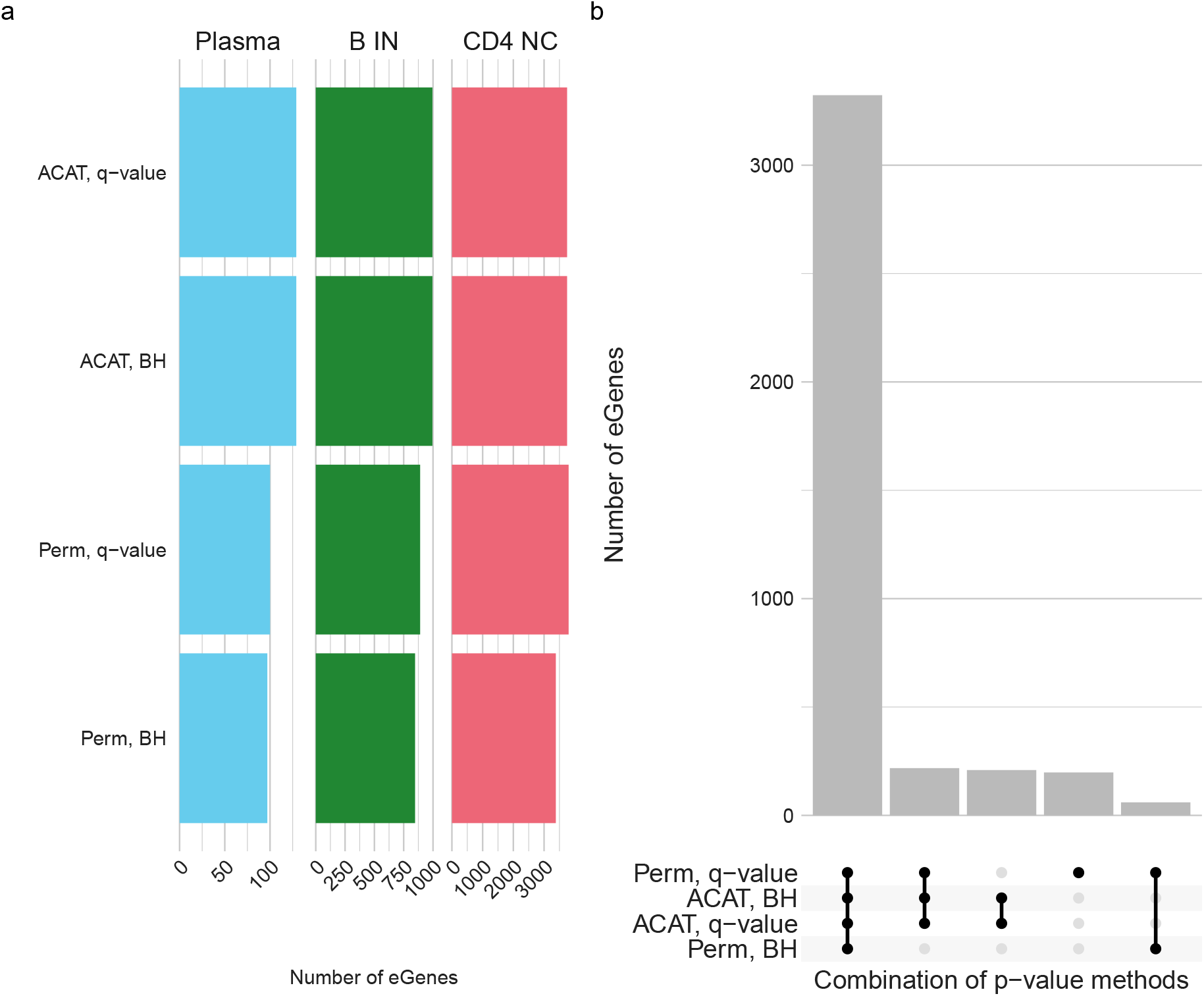
Comparison of different strategies for computing gene-level p-values. a) The number of eGenes called by different methods of computing gene-level p-values at a 5% FDR threshold. b) An upset plot showing the overlap between sets of eGene computed using different methods in the CD4 NC cell type. There are two methodological choices, leading to four combinations of methods. The p-values can be computed using either ACAT or permutations and FDR correction can be performed for using either the Benjamini-Hochberg procedure or with q-values. All results are for tensorQTL computed p-values. The permutation p-values are computed by tensorQTL and the ACAT p-values are computed from the variant-level p-values.

**Supplementary Figure S8:**
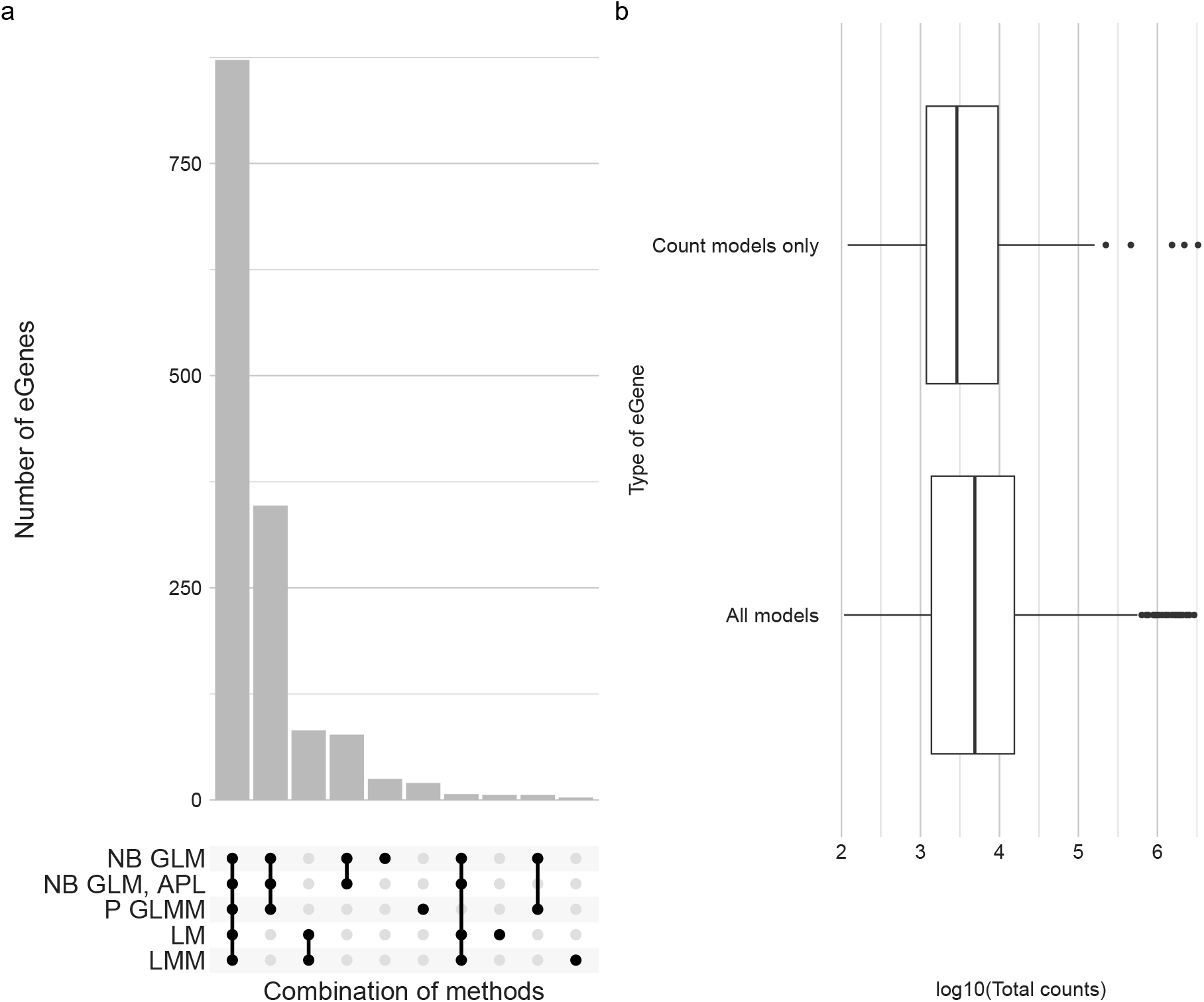
Characteristic of eGenes mapped by quasar. a) Upset plot of eGenes found across quasar methods in the B IN cell type. Most eGenes found are consistent across methods but a minority are found only by the count distribution methods. b) Boxplots of the log_10_ total counts for eGenes found by all methods or by count methods only

