## Supplementary Text for "Flexible and efficient count-distribution and mixed-model methods for eQTL mapping with quasar"

#### Contents

**1 Introduction** **1**

**2 Statistical models for QTL mapping** **1**

**3 Implementation details for quasar** **5**

**4 Score test variance** **13**

**5 Approximating the mixed model score test variance** **14**

#### 1 Introduction

The aim of this supplementary material is to describe the statistical methodology of quasar and justify its particular methodological choices.

#### 2 Statistical models for QTL mapping

In QTL mapping we aim to assess whether genetic variants are statistically associated with the abundance of a feature. The quasar program implements six statistical models for bulk/pseudobulk QTL mapping:

- Linear model (LM),
- Linear mixed model (LMM),

- Poisson generalised linear model (Poisson GLM),
- Negative binomial generalised linear Model (NB GLM),
- Poisson generalised linear mixed model (Poisson GLMM), and
- Negative binomial generalised linear Mixed Model (NB GLMM).

Below we outline the models for a specific feature-variant pair. To perform QTL mapping using the models we test whether the coefficient of genotype, $b$, is zero. Formally, we test the hypothesis $H_0 : b = 0$ against the alternative $H_1 : b \neq 0$.

#### Model notation

Across models we use the following notation:

- $\boldsymbol{X}$ is an $n \times c$ matrix of covariates, such as age, sex and expression and genotype principle components,
- $\boldsymbol{\alpha}$ is a $c$-vector of covariate coefficients,
- $\boldsymbol{g}$ is an $n$-vector of genotype values,
- $b$ is the effect of the genotype on the trait,
- $\boldsymbol{I}_n$ is the $n \times n$ identify matrix, and
- $\boldsymbol{K}$ is an estimated genetic relatedness matrix quantifying genetic similarity between individuals.

#### Linear model

A linear model for QTL mapping is

$$\boldsymbol{y} = \boldsymbol{X\alpha} + \boldsymbol{g}b + \boldsymbol{e} \quad \boldsymbol{e} \sim N(\boldsymbol{0}, \sigma^2 \boldsymbol{I}_n),$$

where

- $\boldsymbol{y}$ is an $n$-vector of the quantitative trait, such as gene expression, appropriately transformed so that it is approximately normally distributed,
- $\boldsymbol{e}$ is an $n$-vector of errors, and
- $\sigma^2$ is a variance parameter.

#### Linear mixed model

A linear mixed model for QTL mapping is

$$\boldsymbol{y} = \boldsymbol{X\alpha} + \boldsymbol{g}b + \boldsymbol{u} + \boldsymbol{e}, \quad \boldsymbol{u} \sim N(\boldsymbol{0}, \sigma_g^2 \boldsymbol{K}), \quad \boldsymbol{e} \sim N(\boldsymbol{0}, \sigma_e^2 \boldsymbol{I}_n),$$

where

- $\boldsymbol{y}$ is an $n$-vector of the quantitative trait, such as gene expression, appropriately transformed so that it is approximately normally distributed,

- $\boldsymbol{u}$ is an $n$-vector of random effects designed to model genetic similarity between individuals,
- $\boldsymbol{e}$ are the residuals errors, and
- $\sigma_g^2$ and $\sigma_e^2$ are the genetic and residual variances, respectively.

The model incorporates a random effect to account for sample relatedness.

#### Poisson generalised linear model

A Poisson generalised linear model for QTL mapping is

$$\boldsymbol{y} \sim \text{Po}(\boldsymbol{\mu}), \quad \log(\boldsymbol{\mu}) = \boldsymbol{X\alpha} + \boldsymbol{g}b + \boldsymbol{o},$$

where

- $\boldsymbol{y}$ is the $n$-vector of feature counts,
- $\boldsymbol{\mu}$ is the mean of $\boldsymbol{y}$, and
- $\boldsymbol{o}$ is a vector of offsets (see Section "Offset Computation" for more information about the offsets used in quasar).

This model directly models the count nature of the data, assuming that $\text{Var}(\boldsymbol{y}) = \boldsymbol{\mu}$.

#### Negative binomial generalised linear model

A negative binomial generalised linear model for QTL mapping is

$$\boldsymbol{y} \sim \text{NB}(\boldsymbol{\mu}, \phi), \quad \log(\boldsymbol{\mu}) = \boldsymbol{X\alpha} + \boldsymbol{g}b + \boldsymbol{o},$$

where

- $\boldsymbol{y}$ is the $n$-vector of feature counts,
- $\boldsymbol{\mu}$ is the mean of $\boldsymbol{y}$,
- $\phi$ is the dispersion parameter of the negative binomial distribution, and
- $\boldsymbol{o}$ is a vector of offsets.

We work with the parameterisation of a negative binomial distribution $W$ where $\mathbf{E}(W) = \mu + \phi\mu^2$. This model accounts for the count nature of the data allowing for overdispersion, enabling it to handle higher variance data than the Poisson model.

#### Poisson generalised linear mixed model

The quasar method also implements a generalised linear mixed model (GLMM) using the Poisson distribution which can be fit to data from $n$ individuals,

$$\boldsymbol{y} \sim \text{Po}(\boldsymbol{\mu}), \quad \log(\boldsymbol{\mu}) = \boldsymbol{X\alpha} + \boldsymbol{g}b + \boldsymbol{u} + \boldsymbol{o} \quad \boldsymbol{u} \sim N(\mathbf{0}, \sigma_g^2 \boldsymbol{K}),$$

where

- $\boldsymbol{y}$ is the $n$-vector of feature counts, and
- $\boldsymbol{u}$ is the random effect.

This model incorporates both sample relatedness and the count nature of the data.

#### Negative binomial generalised linear mixed model

Furthermore, quasar also implements a generalised linear mixed model (GLMM) using the negative binomial distribution which can be fit to data from $n$ individuals,

$$\boldsymbol{y} \sim \mathrm{NB}(\boldsymbol{\mu}, \phi), \quad \log(\boldsymbol{\mu}) = \boldsymbol{X\alpha} + \boldsymbol{g}b + \boldsymbol{u} + \boldsymbol{o}, \quad \boldsymbol{u} \sim N(\boldsymbol{0}, \sigma_g^2 \boldsymbol{K}),$$

where

- $\boldsymbol{y}$ is the $n$-vector of feature counts,
- $\boldsymbol{u}$ is the random effect,
- $\phi$ is the dispersion parameter of the negative binomial distribution, and
- $\boldsymbol{K}$ is the estimated genetic-relatedness matrix.

This model incorporates both sample relatedness and the count nature of the data, while directly accounting for overdispersion in the sampling distribution.

#### Model choice considerations

##### Poisson GLMM vs NB-GLMM

There are strong structural similarities between the PGLMM and NB-GLM models [1]. In particular, they have the same quadratic mean-variance relationship, $\mathrm{Var}(y_i) = \mu_i + c\mu_i^2$. Precisely, for the Poisson GLMM, if we assume that the random effect is $u \sim N(\boldsymbol{0}, \boldsymbol{I})$, i.e., that there is no genetic relatedness between samples, then

$$\boldsymbol{y} \sim \mathrm{Po}(\boldsymbol{\mu}), \quad \log(\boldsymbol{\mu}) = \boldsymbol{X\beta} + \boldsymbol{u}, \quad \boldsymbol{u} \sim N(0, \sigma^2 \boldsymbol{I}_n) \implies \mathrm{Var}(y_i) = \mathbf{E}(y_i) + \exp(\sigma^2 - 1)\mathbf{E}(y_i)^2.$$

On the other hand, in the negative binomial GLM,

$$\boldsymbol{y} \sim \mathrm{NB}(\boldsymbol{\mu}, \phi), \quad \boldsymbol{\mu} = \exp(\boldsymbol{X\beta}) \implies \mathrm{Var}(y_i) = \mathbf{E}(y_i) + \phi\mathbf{E}(y_i)^2.$$

In addition, we can derive under the Negative Binomial GLMM,

$$\boldsymbol{y} \sim \mathrm{NB}(\boldsymbol{\mu}, \phi), \quad \log(\boldsymbol{\mu}) = \boldsymbol{X\beta} + \boldsymbol{u}, \quad \boldsymbol{u} \sim N(0, \sigma^2 \boldsymbol{I}_n),$$

an expression for $\mathrm{Var}(y_i)$. First,

$$\mathbf{E}(y_i) = \mathbf{E}(\mathbf{E}(y_i \mid u_i)) = \mathbf{E}(\exp(\boldsymbol{x}_i^T\boldsymbol{\beta} + u_i)) = \exp(\boldsymbol{x}_i^T\boldsymbol{\beta})M_{u_i}(1) = \exp(\boldsymbol{x}_i^T\boldsymbol{\beta})\exp(\sigma^2/2),$$

where $M_{u_i}$ denotes the moment generating function of $u_i$. Furthermore, the law of total variance, gives that

$$\mathrm{Var}(y_i) = \mathrm{Var}(\mathbf{E}(y_i \mid u_i)) + \mathbf{E}(\mathrm{Var}(y_i \mid u_i)).$$

We can derive each term. First,

$$\begin{aligned}\mathrm{Var}(\mathbf{E}(y_i \mid u_i)) &= \mathrm{Var}(\exp(\boldsymbol{x}_i^T\boldsymbol{\beta} + u_i)) \\ &= \exp(\boldsymbol{x}_i^T\boldsymbol{\beta})^2\mathrm{Var}(\exp(u_i)) \\ &= \exp(\boldsymbol{x}_i^T\boldsymbol{\beta})^2(\exp(2\sigma^2) - \exp(\sigma^2)).\end{aligned}$$

Second,

$$\begin{aligned}\mathbf{E}(\mathrm{Var}(y_i \mid u_i)) &= \mathbf{E}(\mathbf{E}(y_i \mid u_i) + \phi\mathbf{E}(y_i \mid u_i)^2) \\ &= \mathbf{E}(\exp(\boldsymbol{x}_i^T\boldsymbol{\beta} + u_i) + \phi\exp(\boldsymbol{x}_i^T\boldsymbol{\beta} + u_i)^2) \\ &= \exp(\boldsymbol{x}_i^T\boldsymbol{\beta})\exp(\sigma^2/2) + \phi\exp(\boldsymbol{x}_i^T\boldsymbol{\beta})^2\exp(2\sigma^2) \\ &= \mathbf{E}(y_i) + \phi\mathbf{E}(y_i)^2\end{aligned}$$

Combining these expressions gives,

$$\begin{aligned}\mathrm{Var}(y_i) &= \exp(\boldsymbol{x}_i^T\beta)^2(\exp(2\sigma^2) - \exp(\sigma^2)) + \mathbf{E}(y_i) + \phi\mathbf{E}(y_i)^2 \\ &= (\exp(\sigma^2) - 1)(\exp(\boldsymbol{x}_i^T\beta\exp(\sigma^2/2))^2 + \mathbf{E}(y_i) + \phi\mathbf{E}(y_i)^2 \\ &= (\exp(\sigma^2) - 1)(\mathbf{E}(y_i))^2 + \mathbf{E}(y_i) + \phi\mathbf{E}(y_i)^2 \\ &= \mathbf{E}(y_i) + (\phi + \exp(\sigma^2) - 1)\mathbf{E}(y_i)^2.\end{aligned}$$

This expression shows that when the random effects are independent across samples, both the random effect and the negative binomial component of the NB-GLMM induce a quadratic mean variance relationship, and that there is a difficulty in simultaneously identifying $\phi$ and $\sigma^2$ as increasing either one will increase the degree of modelled overdispersion.

In analysis of genetic data, the random effect is constrained to have variance-covariance $\sigma^2\boldsymbol{K}$ where $\boldsymbol{K}$ is the genetic relatedness matrix. If $\boldsymbol{K}$ has strong structure then the $u_i$ values will be constrained to covary, reducing their ability to account for general overdispersion in the data. However, we have also observed that in the common scenario of data from unrelated individuals $\boldsymbol{K} \approx \boldsymbol{I}$, and in this scenario the above results hold approximately and standard algorithms for fitting the NB-GLMM give estimates of $\phi$ very close to 0. This statistical observation also accords with reports that only a P-GLMM is necessary to fit eQTLs [2] and that other attempts to fit the NB-GLMM have used heuristic methods to trade-off between the two sources of overdispersion [3].

### 3 Implementation details for quasar

#### Overall algorithm

The quasar method implements the following algorithm:

1. For each feature we fit the null model with mean model $g(\boldsymbol{y}) = \boldsymbol{X}\boldsymbol{\alpha}$ using one of the six models outlined above.
2. We use the estimated null model to compute a residualised vector of trait values $\tilde{\boldsymbol{y}}$ for each feature. For the linear mixed model we compute

$$\tilde{\boldsymbol{y}} = \hat{\boldsymbol{P}}\boldsymbol{y},$$

where $\boldsymbol{P} = \hat{\boldsymbol{\Sigma}}^{-1} - \hat{\boldsymbol{\Sigma}}^{-1}\boldsymbol{X}(\boldsymbol{X}^T\hat{\boldsymbol{\Sigma}}^{-1}\boldsymbol{X})^{-1}\boldsymbol{X}^T\hat{\boldsymbol{\Sigma}}^{-1}$ and $\hat{\boldsymbol{\Sigma}} = \hat{\sigma}_e^2\boldsymbol{I} + \hat{\sigma}_g^2\boldsymbol{K}$. For the other models we compute

$$\tilde{\boldsymbol{y}} = \frac{d\boldsymbol{\eta}}{d\boldsymbol{\mu}}(\boldsymbol{y} - \hat{\boldsymbol{y}}),$$

where $\boldsymbol{\eta}$ is the linear predictor of the model and $\hat{\boldsymbol{y}}$ are the fitted values from the model. These are the 'working residuals' from the fitted model.

3. Project out the covariates from the genotype vectors. For all $\boldsymbol{g}$ we compute

$$\tilde{\boldsymbol{g}} = \boldsymbol{P_W}\boldsymbol{g} = \boldsymbol{g} - \boldsymbol{X}(\boldsymbol{X}^T\boldsymbol{W}\boldsymbol{X})^{-1}\boldsymbol{X}\boldsymbol{W}\boldsymbol{g},$$

where $\boldsymbol{P_W} = \boldsymbol{I} - (\boldsymbol{X}^T\boldsymbol{W}\boldsymbol{X})^{-1}\boldsymbol{X}^T\boldsymbol{W}$ and $\boldsymbol{W}$ is the diagonal matrix of working weights from the fitted model (see the sections describing the GLM and GLMM fitting algorithms for more information on the working weights). Here and below, for the LMM and LM $\boldsymbol{W} = \boldsymbol{I}$.

4. To test for an effect we use the score statistic:

$$Z = \frac{\tilde{\boldsymbol{g}}^T\boldsymbol{W}\tilde{\boldsymbol{y}}}{V}, \quad V^2 = \begin{cases} \hat{\sigma}^2\tilde{\boldsymbol{g}}\boldsymbol{W}\tilde{\boldsymbol{g}}, & \text{LMM} \\ \hat{r}\tilde{\boldsymbol{g}}\boldsymbol{W}\tilde{\boldsymbol{g}}, & \text{P-GLMM, NB-GLMM} \\ \tilde{\boldsymbol{g}}\boldsymbol{W}\tilde{\boldsymbol{g}}, & \text{LM, GLM} \end{cases}$$

where,

$$\hat{\sigma}^2 = \frac{\tilde{\boldsymbol{y}}^T\tilde{\boldsymbol{y}}}{n-c}, \text{ and } \hat{r} = \frac{\text{tr}(\boldsymbol{P})}{\text{tr}(\boldsymbol{W}\boldsymbol{P_W})} - \frac{2\text{tr}(\boldsymbol{P}\boldsymbol{W}\boldsymbol{P_W})}{\text{tr}(\boldsymbol{W}\boldsymbol{P_W})^2} + \frac{2\text{tr}(\boldsymbol{W}\boldsymbol{P_W}\boldsymbol{W}\boldsymbol{P_W})\text{tr}(\boldsymbol{P})}{\text{tr}(\boldsymbol{W}\boldsymbol{P_W})^3},$$

where $\boldsymbol{P} = \hat{\boldsymbol{\Sigma}}^{-1} - \hat{\boldsymbol{\Sigma}}^{-1}\boldsymbol{X}(\boldsymbol{X}^T\hat{\boldsymbol{\Sigma}}^{-1}\boldsymbol{X})\boldsymbol{X}^T\hat{\boldsymbol{\Sigma}}^{-1}$, $\hat{\boldsymbol{\Sigma}} = \boldsymbol{W}^{-1} + \hat{\sigma}^2\boldsymbol{K}$ and $\boldsymbol{P_W} = \boldsymbol{I} - (\boldsymbol{X}^T\boldsymbol{W}\boldsymbol{X})^{-1}\boldsymbol{X}^T\boldsymbol{W}$. To compute a $p$-value we use the asymptotic approximation $Z \sim N(0,1)$ under $H_0$. We also output the estimated effect size $\hat{b}$ and its standard error associated with this score test, calculated as

$$\hat{b} = \frac{\tilde{\boldsymbol{g}}^T\boldsymbol{W}\tilde{\boldsymbol{y}}}{V^2}, \quad \text{se}(\hat{b}) = \frac{1}{V}.$$

#### LM fitting algorithm

We use the standard analytic formula, $\hat{\boldsymbol{\beta}} = (\boldsymbol{X}^T\boldsymbol{X})^{-1}\boldsymbol{X}^T\boldsymbol{y}$ to fit the linear model.

#### LMM fitting algorithm

We use the efficient algorithm presented by Lippert et al [4] to fit the LMM, which is outlined below. We fit the 'null' LMM without the genotype so we have the model,

$$\boldsymbol{y} \sim N(\boldsymbol{X}\boldsymbol{\alpha}, \sigma_g^2\boldsymbol{K} + \sigma_e^2\boldsymbol{I}_n).$$

Let and $\boldsymbol{\Sigma} = \sigma_g^2\boldsymbol{K} + \sigma_e^2\boldsymbol{I}$. The REML log-likelihood is then

$$l_{\text{REML}}(\sigma_g^2, \sigma_e^2, \boldsymbol{\beta}) = -\frac{1}{2}\log|\boldsymbol{\Sigma}| - \frac{1}{2}\log|\boldsymbol{X}^T\boldsymbol{\Sigma}^{-1}\boldsymbol{X}| - \frac{1}{2}(\boldsymbol{y} - \boldsymbol{X}\boldsymbol{\beta})^T\boldsymbol{\Sigma}^{-1}(\boldsymbol{y} - \boldsymbol{X}\boldsymbol{\beta}).$$

We re-parametrise $\boldsymbol{\Sigma} = \sigma_g^2(\boldsymbol{K} + \delta\boldsymbol{I})$, where $\delta = \sigma_e^2/\sigma_g^2$ and decompose $\boldsymbol{K} = \boldsymbol{Q}\boldsymbol{\Lambda}\boldsymbol{Q}^T$, where $\boldsymbol{Q}$ is the orthogonal matrix of eigenvectors and $\boldsymbol{\Lambda}$ is a diagonal matrix of eigenvalues. Then

$$\boldsymbol{\Sigma} = \sigma_g^2(\boldsymbol{K} + \delta\boldsymbol{I}) = \sigma_g^2(\boldsymbol{Q}\boldsymbol{\Lambda}\boldsymbol{Q}^T + \delta\boldsymbol{I}) = \sigma_g^2\boldsymbol{Q}(\boldsymbol{\Lambda} + \delta\boldsymbol{I})\boldsymbol{Q}^T = \sigma_g^2\boldsymbol{Q}\boldsymbol{D}_\delta\boldsymbol{Q}^T,$$

where we define $\boldsymbol{D}_\delta = \boldsymbol{\Lambda} + \delta \boldsymbol{I}$. Therefore,

$$\boldsymbol{\Sigma}^{-1} = \frac{1}{\sigma_g^2}\boldsymbol{Q}(\boldsymbol{\Lambda}+\delta\boldsymbol{I})^{-1}\boldsymbol{Q}^T = \frac{1}{\sigma_g^2}\boldsymbol{Q}\boldsymbol{D}_\delta^{-1}\boldsymbol{Q}^T,$$

and

$$\log|\boldsymbol{\Sigma}| = n\log\sigma_g^2 + \log|\boldsymbol{\Lambda}+\delta\boldsymbol{I}_n| = n\log\sigma_g^2 + \log|\boldsymbol{D}_\delta|.$$

Using these expressions we rewrite the REML log-likelihood as

$$\begin{aligned} l_{\text{REML}}(\sigma_g^2,\delta,\boldsymbol{\beta}) &= -\frac{n}{2}\log\sigma_g^2 - \frac{1}{2}\log|\boldsymbol{D}_\delta| - \frac{1}{2}\log|\boldsymbol{X}^T\frac{1}{\sigma_g^2}\boldsymbol{Q}\boldsymbol{D}_\delta^{-1}\boldsymbol{Q}^T\boldsymbol{X}| \\ &\quad - \frac{1}{2}(\boldsymbol{y}-\boldsymbol{X}\boldsymbol{\beta})^T\frac{1}{\sigma_g^2}\boldsymbol{Q}\boldsymbol{D}_\delta^{-1}\boldsymbol{Q}^T(\boldsymbol{y}-\boldsymbol{X}\boldsymbol{\beta}^T) \\ &= -\frac{n-\nu}{2}\log\sigma_g^2 - \frac{1}{2}\log|\boldsymbol{D}_\delta| - \frac{1}{2}\log|\tilde{\boldsymbol{X}}^T\boldsymbol{D}_\delta^{-1}\tilde{\boldsymbol{X}}| \\ &\quad - \frac{1}{2\sigma_g^2}(\tilde{\boldsymbol{y}}-\tilde{\boldsymbol{X}}\boldsymbol{\beta})^T\boldsymbol{D}_\delta^{-1}(\tilde{\boldsymbol{y}}-\tilde{\boldsymbol{X}}\boldsymbol{\beta}), \end{aligned}$$

where $\tilde{\boldsymbol{X}} = \boldsymbol{Q}^T\boldsymbol{X}$, $\tilde{\boldsymbol{y}} = \boldsymbol{Q}^T\boldsymbol{y}$ and $\nu = c+1$. Differentiating this likelihood with respect to $\boldsymbol{\beta}$ and $\sigma$ gives closed forms for the (restricted) maximum likelihood estimators for these parameters as functions of $\delta$,

$$\hat{\boldsymbol{\beta}}(\delta) = (\tilde{\boldsymbol{X}}^T\boldsymbol{D}_\delta^{-1}\tilde{\boldsymbol{X}})^{-1}\tilde{\boldsymbol{X}}^T\boldsymbol{D}_\delta^{-1}\tilde{\boldsymbol{y}}$$

and

$$\hat{\sigma}_g(\delta,\boldsymbol{\beta}) = \frac{1}{n-\nu}(\tilde{\boldsymbol{y}}-\tilde{\boldsymbol{X}}\boldsymbol{\beta})^T\boldsymbol{D}_\delta^{-1}(\tilde{\boldsymbol{y}}-\tilde{\boldsymbol{X}}\boldsymbol{\beta}).$$

Plugging in $\hat{\boldsymbol{\beta}}$ gives

$$\hat{\sigma}^2_{\boldsymbol{\beta}=\hat{\boldsymbol{\beta}}}(\delta) = \frac{1}{n-\nu}\boldsymbol{y}^T\boldsymbol{A}_\delta\boldsymbol{y},$$

where $\boldsymbol{A}_\delta = \boldsymbol{D}_\delta^{-1} - \boldsymbol{D}_\delta^{-1}\tilde{\boldsymbol{X}}(\tilde{\boldsymbol{X}}^T\boldsymbol{D}_\delta^{-1}\tilde{\boldsymbol{X}})^{-1}\tilde{\boldsymbol{X}}\boldsymbol{D}_\delta^{-1}$. Plugging these expressions into the REML log-likelihood we obtain a profile log-likelihood that is a function of only $\delta$:

$$\begin{aligned} l_{\text{REML}}(\delta) &= l_{\text{REML}}(\hat{\sigma}_g^2,\delta,\hat{\boldsymbol{\alpha}}) \\ &= -\frac{n-\nu}{2}\log\hat{\sigma}_g^2 - \frac{1}{2}\log|\boldsymbol{D}_\delta| - \frac{1}{2}\log|\tilde{\boldsymbol{X}}\boldsymbol{D}_\delta^{-1}\tilde{\boldsymbol{X}}| - \frac{1}{2\hat{\sigma}_g^2}(\tilde{\boldsymbol{y}}-\tilde{\boldsymbol{X}}\hat{\boldsymbol{\beta}})^T\boldsymbol{D}_\delta^{-1}(\tilde{\boldsymbol{y}}-\tilde{\boldsymbol{X}}\hat{\boldsymbol{\beta}}) \\ &= -\frac{n-\nu}{2}\log\boldsymbol{y}^T\boldsymbol{A}_\delta\boldsymbol{y}^T - \frac{1}{2}\log|\boldsymbol{D}_\delta| - \frac{1}{2}\log|\tilde{\boldsymbol{X}}\boldsymbol{D}_\delta^{-1}\tilde{\boldsymbol{X}}| + c, \end{aligned}$$

where $c$ is a constant. We optimize this one-dimensional objective using the Brent algorithm as in [5, 6].

#### GLM fitting algorithm

To fit GLMs we use the classical Iteratively Reweighted Weighted Least Squares (IRWLS) algorithm. We assume we have a link function $g(\boldsymbol{\mu})$, the inverse link function $h(\boldsymbol{\eta}) = g^{-1}(\boldsymbol{\eta})$ and variance function $v(\boldsymbol{\mu})$. The fitting algorithm is then:

1. Initialise $\boldsymbol{\mu}^0$ using $\boldsymbol{y}$,

2. At the $t$-th iteration, until convergence, form the working weights

$$\boldsymbol{W}^{(t)} = \text{diag}(\boldsymbol{w}), \quad \boldsymbol{W}_{ii} = \frac{1}{v(\boldsymbol{\mu}_i)g'(\boldsymbol{\mu}_i)^2},$$

3. Compute the working vector,

$$\tilde{\boldsymbol{y}}^{(t)} = \boldsymbol{\eta} + g'(\boldsymbol{\mu})(\boldsymbol{y} - \boldsymbol{\mu}),$$

4. Solve the weighted least squares problem,

$$\boldsymbol{\beta}^{(t+1)} = (\boldsymbol{X}^T\boldsymbol{W}^{(t)}\boldsymbol{X})^{-1}\boldsymbol{X}^T\boldsymbol{W}^{(t)}\tilde{\boldsymbol{y}}^{(t)}$$

, and

5. Set $\boldsymbol{\eta} = \boldsymbol{X}\boldsymbol{\beta}^{(t+1)}$ $\boldsymbol{\mu}^{(t+1)} = h(\boldsymbol{\eta}^{(t+1)})$.

In the Poisson GLM, we use $g(\boldsymbol{\mu}) = \log(\boldsymbol{\mu})$, $v(\boldsymbol{\mu}) = \boldsymbol{\mu}$ and initialise $\boldsymbol{\mu} = \boldsymbol{y} + 0.1$, following the glm function in R. For the negative binomial GLM where $\phi$ is known, we use $g(\boldsymbol{\mu}) = \log(\boldsymbol{\mu})$, $v(\boldsymbol{\mu}) = \boldsymbol{\mu} + \phi\boldsymbol{\mu}^2$ and we initialise $\boldsymbol{\mu} = \boldsymbol{y} + 0.1$.

#### Negative binomial GLM estimation algorithm

To fit the negative binomial GLM when $\phi$ is unknown we use the maximum likelihood algorithm from `MASS::glm.nb()`. Specifically, we use an algorithm that alternates between estimating $\phi$ and fitting a negative binomial model with known $\phi$. The algorithm is:

1. Fit a Poisson GLM to produce fitted values $\hat{\boldsymbol{\mu}}$. Then, until convergence, repeat the following two steps.
2. Estimate $\hat{\phi}$ based on $\hat{\boldsymbol{\mu}}$ and $\boldsymbol{y}$.
3. Fit a negative binomial GLM with $\phi$ fixed to the estimated value.

#### Negative binomial dispersion estimation

We implement two approaches for estimating the negative binominal dispersion parameter in quasar, a maximum-likelihood approach, taken from `MASS::theta.ml()` and an approach based on the Cox-Reid adjusted profile likelihood [7, 8], a key component of the edgeR and DESeq2 methods for differential expression testing with RNA-seq data.

##### Maximum likelihood

Following `MASS::theta.ml()` we maximise the log-likelihood as function of $\phi$ using a Newton-Raphson scheme.

##### Adjusted profile likelihood

We maximise the adjusted profile likelihood,

$$\text{APL}(\phi) = l(\phi) - \frac{1}{2}\log|\boldsymbol{X}^T\boldsymbol{W}\boldsymbol{X}|,$$

where $l(\phi)$ is the usual log likelihood , $|\cdot|$ denotes the determinant, $\boldsymbol{X}^T\boldsymbol{W}\boldsymbol{X}$ is the Fisher matrix, where $\boldsymbol{X}$ is the matrix of covariates and $\boldsymbol{W}$ is the diagonal matrix of working weights. We maximise APL($\phi$) using Brent's algorithm, which is chosen for convenience as it is also used to fit the LMM.

#### GLMM fitting algorithm

We use the penalised quasi-likelihood (PQL) algorithm to fit the GLMM. The PQL algorithm has previously been used to fit logistic linear mixed models in the context of GWAS analysis of binary traits [9] and is also used to fit a Poisson Mixed model to scRNA-seq data by SAIGE-QTL [10]. We outline the algorithm for fitting a Poisson GLMM, outlined above. We consider fitting the model under the null where $b = 0$ and for notational convenience let $\boldsymbol{R} = \sigma^2\boldsymbol{K}$. We approximate the conditional log likelihood for the $i$th individual $l_i(\boldsymbol{\alpha}, \sigma^2, b \mid \boldsymbol{u})$ by the log quasi-likelihood $ql_i(\boldsymbol{\alpha}, \sigma^2, b \mid \boldsymbol{u})$ defined by

$$ql_i(\boldsymbol{\alpha}, \sigma^2 \mid \boldsymbol{u}) = \int_{y_i}^{\mu_i} \frac{y_i - \mu}{\mu} d\mu.$$

The joint quasi-likelihood is then

$$\begin{aligned} l(\boldsymbol{\alpha}, \sigma^2) &= \int \exp\left(\sum_{i=1}^{n} ql_i(\boldsymbol{\alpha})\right) p(\boldsymbol{u} \mid \boldsymbol{\tau}) d\boldsymbol{u} \\ &= \int \exp\left(\sum_{i=1}^{n} ql_i(\boldsymbol{\alpha})\right) (2\pi)^{-\frac{N}{2}} |\boldsymbol{R}|^{-\frac{1}{2}} \exp\left(\boldsymbol{u}^T\boldsymbol{R}^{-1}\boldsymbol{u}\right) d\boldsymbol{u} \\ &= (2\pi)^{-\frac{N}{2}} |\boldsymbol{R}|^{-\frac{1}{2}} \int \exp\left(\sum_{i=1}^{n} ql_{i,j}(\boldsymbol{\alpha}) - \boldsymbol{u}^T\boldsymbol{R}^{-1}\boldsymbol{u}\right). \end{aligned}$$

We approximate the integral above using the Laplace approximation. Let

$$h(\boldsymbol{u}) = \sum_{i=1}^{n} ql_i(\boldsymbol{\alpha}) - \boldsymbol{u}^T\boldsymbol{R}^{-1}\boldsymbol{u}.$$

Then the Laplace approximation is

$$\int \exp(h(\boldsymbol{u})) \approx (2\pi)^{\frac{N}{2}} |-h''(\boldsymbol{u}_0)|^{-\frac{1}{2}} \exp(h(\boldsymbol{u}_0)),$$

where $\boldsymbol{u}_0$ is the solution to $h'(\boldsymbol{u}) = 0$. We have that

$$h'(\boldsymbol{u}) = \sum_{i=1}^{n} \frac{\partial ql_i(\boldsymbol{\alpha}, \sigma^2, \mid \boldsymbol{u})}{\partial \boldsymbol{u}} - \boldsymbol{u}^T\boldsymbol{R}^{-1}$$

and

$$\begin{aligned} h''(\boldsymbol{u}) &= \sum_{i=1}^{n} \frac{\partial^2 ql_{i,j}(\boldsymbol{\alpha}, \sigma^2 \mid \boldsymbol{u})}{\partial \boldsymbol{u} \partial \boldsymbol{u}^T} - \boldsymbol{R}^{-1} \\ &= \boldsymbol{W} + \boldsymbol{R}^{-1}. \end{aligned}$$

Plugging these expressions into the Laplace approximation and taking the log gives the following expression for the approximate joint log quasi-likelihood $l(\boldsymbol{\alpha}, \sigma^2)$ up to a constant,

$$
\begin{aligned}
l(\boldsymbol{\alpha}, \sigma^2) &= -\frac{1}{2}\log|\boldsymbol{R}| - \frac{1}{2}\log\left|\sum_{i=1}^{n}\mu_i + \boldsymbol{R}^{-1}\right| + \sum_{i=1}^{n} ql_i(\boldsymbol{\alpha}, \boldsymbol{u}_0) - \boldsymbol{u}_0^T\boldsymbol{R}^{-1}\boldsymbol{u}_0 \\
&= -\frac{1}{2}\log|\boldsymbol{R}\boldsymbol{W} + \boldsymbol{I}| + \sum_{i=1}^{n} ql_{i,j}(\boldsymbol{\alpha} \mid \boldsymbol{u}_0) - \boldsymbol{u}_0^T\boldsymbol{R}^{-1}\boldsymbol{u}_0,
\end{aligned}
$$

where $\boldsymbol{W} = \text{diag}(\mu_i)$ is an $n \times n$ diagonal matrix.

##### Estimation of fixed and random effect

First, we obtain estimates of $\boldsymbol{\alpha}$ and $\boldsymbol{u}$ conditional on the variance estimates. We can compute the score equations as

$$
\frac{\partial ql(\boldsymbol{\alpha}, \sigma^2)}{\partial \boldsymbol{\alpha}} = \sum_{i=1}^{n} \frac{\partial ql_i(\boldsymbol{\alpha} \mid \boldsymbol{u}_0)}{\partial \mu_{i,j}} \frac{\partial \mu_i}{\eta_i} \frac{\partial \mu_i}{\partial \boldsymbol{\alpha}} = \sum_{i=1}^{n} \frac{y_i - \mu_i}{\mu_i} \times \mu_i \times \boldsymbol{X}_i = \boldsymbol{X}^T(\boldsymbol{y} - \boldsymbol{\mu}),
$$

and

$$
\frac{\partial ql(\boldsymbol{\alpha}, \sigma^2)}{\partial \boldsymbol{u}} = \boldsymbol{y} - \boldsymbol{\mu} - \boldsymbol{R}^{-1}\boldsymbol{u}.
$$

We define the working pseudo-data $\tilde{\boldsymbol{y}} = \boldsymbol{\eta} + \boldsymbol{W}(\boldsymbol{y} - \boldsymbol{\mu})$ where $\boldsymbol{\eta} = (\eta_1, \ldots, \eta_n)$ and $\boldsymbol{\mu} = (\mu_1, \ldots \mu_n)$. Then $\boldsymbol{y} - \boldsymbol{\mu} = \boldsymbol{W}(\tilde{\boldsymbol{y}} - \boldsymbol{X}\boldsymbol{\alpha} - \boldsymbol{u})$. Substituting this expression into the score function allows us to write the score equations as

$$
\begin{pmatrix} \boldsymbol{X}^T\boldsymbol{W}\boldsymbol{X} & \boldsymbol{X}^T\boldsymbol{W} \\ \boldsymbol{W}\boldsymbol{X} & \boldsymbol{W} + \boldsymbol{R} \end{pmatrix} \begin{pmatrix} \boldsymbol{\alpha} \\ \boldsymbol{u} \end{pmatrix} = \begin{pmatrix} \boldsymbol{X}^T\boldsymbol{W}\tilde{\boldsymbol{y}} \\ \boldsymbol{W}\tilde{\boldsymbol{y}} \end{pmatrix}.
$$

Solving the score equations using the gives the updates

$$
\begin{aligned}
\hat{\boldsymbol{\beta}} &= (\boldsymbol{X}^T\boldsymbol{\Sigma}^{-1}\boldsymbol{X})^{-1}\boldsymbol{X}\boldsymbol{\Sigma}^{-1}\tilde{\boldsymbol{y}}, \\
\hat{\boldsymbol{u}} &= \boldsymbol{R}\,\boldsymbol{\Sigma}^{-1}(\tilde{\boldsymbol{y}} - \boldsymbol{X}\hat{\boldsymbol{\beta}}),
\end{aligned}
$$

where $\boldsymbol{\Sigma} = \boldsymbol{W}^{-1} + \boldsymbol{R}$.

##### Estimation of variance components

Next, given $\hat{\boldsymbol{\alpha}}$, $\hat{\boldsymbol{u}}$, the log likelihood of the variance component can be derived as

$$
ql(\hat{\boldsymbol{\alpha}}, \sigma^2) = c - \frac{1}{2}\log|\boldsymbol{\Sigma}| - \tilde{\boldsymbol{y}}^T\boldsymbol{P}\tilde{\boldsymbol{y}},
$$

where $\boldsymbol{P} = \boldsymbol{\Sigma}^{-1} - \boldsymbol{\Sigma}^{-1}\boldsymbol{X}(\boldsymbol{X}^T\boldsymbol{\Sigma}^{-1})\boldsymbol{X}^T\boldsymbol{\Sigma}^{-1}$, $\boldsymbol{\Sigma} = \boldsymbol{W}^{-1} + \boldsymbol{R}$ and $c$ is a constant. We instead maximise the corresponding REML,

$$
ql_R(\hat{\boldsymbol{\alpha}}(\sigma^2), \sigma^2) = c - \frac{1}{2}\log|\Sigma| - \frac{1}{2}\log|\boldsymbol{X}^T\boldsymbol{\Sigma}^{-1}\boldsymbol{X}| - \frac{1}{2}\tilde{\boldsymbol{y}}^T\boldsymbol{P}\tilde{\boldsymbol{y}}.
$$

The score function for $\sigma^2$ is, $\boldsymbol{U}_{\sigma^2}$:

$$
\boldsymbol{U}_{\sigma^2} = \frac{\partial ql_R(\hat{\boldsymbol{\alpha}}(\sigma^2), \sigma^2)}{\partial \sigma^2} = \frac{1}{2}\left(\boldsymbol{Y}^T\boldsymbol{P}\boldsymbol{K}\boldsymbol{P}\boldsymbol{Y} - \text{tr}(\boldsymbol{P}\boldsymbol{K})\right),
$$

and the corresponding observed and expected informations are:

$$\boldsymbol{J}_{\sigma^2} = -\frac{\partial ql_R(\boldsymbol{\alpha}(\sigma^2), \sigma^2}{\partial\sigma^2)} = -\frac{1}{2}\text{tr}(\boldsymbol{PKPV}_k) + \tilde{\boldsymbol{y}}^T\boldsymbol{PKPKP}\tilde{\boldsymbol{y}},$$

$$\boldsymbol{E}(\boldsymbol{J}_\tau) = \boldsymbol{E}\left(-\frac{\partial ql_R(\boldsymbol{\alpha}(\sigma^2), \sigma^2)}{\partial\sigma^2}\right) = -\frac{1}{2}\text{tr}(\boldsymbol{PKPK})$$

Both the observed and expected information contain traces of large, expensive-to-compute matrices. To avoid the need for these computations, we use the average information, as in the Average Information REML (AI REML) algorithm. The average information is then,

$$\text{AI} = \frac{1}{2}\tilde{\boldsymbol{y}}^T\boldsymbol{PKPKP}\tilde{\boldsymbol{y}}.$$

**Overall algorithm**

The overall algorithm for fitting a null GLMM is thus:

1. Fit a Poisson GLM to get an initial estimates of $\hat{\boldsymbol{\beta}}_{(0)}$ and working vector $\boldsymbol{\mu}_{(0)}$.
2. At the *ith* iteration: update

$$\boldsymbol{\Sigma} = \boldsymbol{W}^{-1} + \boldsymbol{R}, \quad \boldsymbol{P} = \boldsymbol{\Sigma}^{-1} - \boldsymbol{\Sigma}^{-1}\boldsymbol{X}(\boldsymbol{X}^T\boldsymbol{\Sigma}^{-1}\boldsymbol{X})\boldsymbol{X}^T\boldsymbol{\Sigma}^{-1}.$$

3. Update $\hat{\sigma}^2$ as

$$\hat{\sigma}^2_{(i)} = \hat{\sigma}^2_{(i-1)} + \alpha\left(\text{AI}\Big|_{\sigma^2_{(i-1)}}\right)^{-1}\boldsymbol{U}_{\hat{\sigma^2}_{(i-1)}},$$

   where $\alpha$ is a step-size parameter.
4. Update $\boldsymbol{\beta}$, $\boldsymbol{u}$, using $\tilde{\boldsymbol{y}}$, $\hat{\sigma^2}_{(i)}$, as

$$\hat{\boldsymbol{\beta}} = (\boldsymbol{X}^T\boldsymbol{\Sigma}^{-1}\boldsymbol{X})^{-1}\boldsymbol{X}\boldsymbol{\Sigma}^{-1}\tilde{\boldsymbol{y}},$$
$$\hat{\boldsymbol{u}} = \boldsymbol{R}\,\boldsymbol{\Sigma}^{-1}(\tilde{\boldsymbol{y}} - \boldsymbol{X}\hat{\boldsymbol{\beta}}).$$

5. Update $\tilde{\boldsymbol{y}}$ using $\hat{\boldsymbol{\alpha}}_{(i)}$, $\hat{\boldsymbol{u}}_{(i)}$: $\hat{\boldsymbol{\tau}}_{(i)}$

$$\tilde{\boldsymbol{y}} = \boldsymbol{\eta} + \boldsymbol{W}(\boldsymbol{y} - \boldsymbol{\mu}).$$

6. Repeat steps 2-4 until

$$\max\left(\frac{|\hat{\boldsymbol{\beta}}_{(i)} - \hat{\boldsymbol{\beta}}_{(i-1)}|}{|\boldsymbol{\beta}_{(i)}| + |\hat{\boldsymbol{\beta}}_{(i-1)}|}, \frac{|\hat{\sigma}^2_{(i)} - \hat{\sigma}^2_{(i-1)}|}{|\hat{\sigma}_{(i)}| + |\hat{\sigma}^2_{(i-1)}|}\right) < \delta,$$

   where $\delta$ is pre-specified tolerance.

##### Additional implementation details

The implementation of this algorithm in quasar includes several computational optimizations. First, inspired by SAIGE-QTL, we fit a GLM to get initial estimates of $\boldsymbol{\beta}$, $\boldsymbol{\mu}$. In practice, we found that this approach led to more stable inference. Second, following the PQLSeq2 GLMM implementation we add a step-size parameter to the update of $\sigma^2$. This parameter is initially set to 1 and decreased by a multiplicative factor 0.9 every 10 iterations. In addition, if the update to $\sigma^2$ would lead to it being negative we halve the step-size until the update does not cause $\sigma^2 < 0$. Finally, we use LDL decompositions of $\boldsymbol{\Sigma}^{-1}$ and $\boldsymbol{X}^T\boldsymbol{\Sigma}^{-1}\boldsymbol{X}$ to avoid explicit computation of their inverses in the algorithm.

##### Negative Binomial GLMM fitting algorithm

To fit the Negative Binomial GLMM we use the same algorithm used for the negative binominal GLM, replacing the Poisson GLM with a Poisson GLMM. The dispersion parameter can be estimated using either maximum likelihood or adjusted profile likelihood. Note that due to structural similarities between the Poisson GLMM and Negative Binomial GLM the Negative Binominal GLMM $\sigma^2$ and $\phi$ parameters are difficult to jointly identify if the GRM is not highly structured.

#### Offset computation

The quasar program computes the sum of the counts across genes as the offset for a particular sample. That is, if $y_{i,j}$ denotes the count for sample $i$ and gene $j$, the offset for sample $i$, $o_i$ is defined as

$$o_i = \sum_{j=1}^{G} y_{i,j},$$

where $G$ is the number of measured genes. In future, other methods of estimating the offset and the ability to pass user-computed offsets may be implemented.

#### Residualisation computation

When performing residualisation using the linear mixed model we wish to compute $\boldsymbol{P}\boldsymbol{y}$ where $\boldsymbol{P} = \boldsymbol{\Sigma}^{-1} - \boldsymbol{\Sigma}^{-1}\boldsymbol{X}(\boldsymbol{X}^T\boldsymbol{\Sigma}^{-1}\boldsymbol{X})^{-1}\boldsymbol{X}^T\boldsymbol{\Sigma}^{-1}$. (For notational convenience we suppress the fact that $\boldsymbol{\Sigma}$ is estimated.) Recall that, $\boldsymbol{\Sigma}^{-1} = \frac{1}{\sigma^2}\boldsymbol{Q}\boldsymbol{D}^{-1}\boldsymbol{Q}^T$, $\tilde{\boldsymbol{y}} = \boldsymbol{Q}^T\boldsymbol{y}$ $\tilde{\boldsymbol{X}} = \boldsymbol{Q}^T\boldsymbol{X}$, and $\hat{\boldsymbol{\beta}} = (\boldsymbol{X}^T\boldsymbol{\Sigma}^{-1}\boldsymbol{X})^{-1}\boldsymbol{X}^T\boldsymbol{\Sigma}^{-1}\boldsymbol{y}$. Then we have,

$$\begin{aligned}
\boldsymbol{P}\boldsymbol{y} &= (\boldsymbol{\Sigma}^{-1} - \boldsymbol{\Sigma}^{-1}\boldsymbol{X}(\boldsymbol{X}^T\boldsymbol{\Sigma}^{-1}\boldsymbol{X})^{-1}\boldsymbol{X}^T\boldsymbol{\Sigma}^{-1})\boldsymbol{y} \\
&= \boldsymbol{\Sigma}^{-1}\boldsymbol{y} - \boldsymbol{\Sigma}^{-1}\boldsymbol{X}\hat{\boldsymbol{\beta}} \\
&= \frac{1}{\sigma^2}\boldsymbol{Q}\boldsymbol{D}^{-1}\boldsymbol{Q}^T\boldsymbol{y} - \frac{1}{\sigma^2}\boldsymbol{Q}\boldsymbol{D}^{-1}\boldsymbol{Q}^T\boldsymbol{X}\hat{\boldsymbol{\beta}} \\
&= \frac{1}{\sigma^2}\boldsymbol{Q}\boldsymbol{D}^{-1}(\tilde{\boldsymbol{y}} - \tilde{\boldsymbol{X}}\hat{\boldsymbol{\beta}}).
\end{aligned}$$

### 4 Score test variance

#### Linear model and generalised linear model

As in Generalized Linear Models With Examples in R (page 272) [11], the score statistic for including $\boldsymbol{g}$ into the GLM with mean model $\boldsymbol{y} = \boldsymbol{X\alpha}$ is

$$Z = \frac{\tilde{\boldsymbol{g}}^T \boldsymbol{W} \boldsymbol{r}}{(\tilde{\boldsymbol{g}}^T \boldsymbol{W} \tilde{\boldsymbol{g}})^{1/2}},$$

where $\boldsymbol{r}$ are the working residuals of the fitted model, $\boldsymbol{W}$ is the diagonal matrix of working weights from the fitted model and

$$\tilde{\boldsymbol{g}} = \boldsymbol{g} - \boldsymbol{X}(\boldsymbol{X}^T \boldsymbol{W} \boldsymbol{X})^{-1} \boldsymbol{X}^T \boldsymbol{W} \boldsymbol{g},$$

the residuals from projecting out the covariates currently in the model. The linear model is a special case of the GLM with $\boldsymbol{W} = \boldsymbol{I}$.

#### Linear mixed model

Under the linear mixed model we have that

$$U = \frac{d\, l_{\mathrm{REML}}}{db} = \boldsymbol{g}^T \boldsymbol{P} \boldsymbol{y}, \quad V = \mathrm{Var}_{H_0}(U) = \boldsymbol{g}^T \boldsymbol{P} \boldsymbol{g}$$

evaluated at the values of the MLEs, $\hat{\boldsymbol{\alpha}}, \hat{\sigma}_e^2, \hat{\sigma}_g^2$, where $\boldsymbol{P} = \hat{\boldsymbol{\Sigma}}^{-1} - \hat{\boldsymbol{\Sigma}}^{-1} \boldsymbol{X}(\boldsymbol{X}^T \hat{\boldsymbol{\Sigma}}^{-1} \boldsymbol{X})^{-1} \boldsymbol{X}^T \hat{\boldsymbol{\Sigma}}^{-1}$ and $\hat{\boldsymbol{\alpha}} = (\boldsymbol{X}^T \hat{\boldsymbol{\Sigma}}^{-1} \boldsymbol{X})^{-1} \boldsymbol{X}^T \hat{\boldsymbol{\Sigma}}^{-1} \boldsymbol{y}$. These expressions can be derived by taking the derivative of $l_{\mathrm{REML}}$ with respect to $b$,

$$\begin{aligned} \frac{d\, l_{\mathrm{REML}}}{db} &= \frac{d}{db}\left(-\frac{1}{2}(\boldsymbol{y} - \boldsymbol{X\alpha} - \boldsymbol{g}b)^T \boldsymbol{\Sigma}^{-1} (\boldsymbol{y} - \boldsymbol{X\alpha} - \boldsymbol{g}b)\right) \\ &= \frac{d}{db}\left(b\boldsymbol{g}^T \boldsymbol{\Sigma}^{-1} \boldsymbol{y} - b\boldsymbol{g}^T \boldsymbol{\Sigma}^{-1} \boldsymbol{X\alpha} - \frac{1}{2} b^2 \boldsymbol{g}^T \boldsymbol{\Sigma}^{-1} \boldsymbol{g}\right) \\ &= \boldsymbol{g}^T \boldsymbol{\Sigma}^{-1} \boldsymbol{y} - \boldsymbol{g}^T \boldsymbol{\Sigma}^{-1} \boldsymbol{X\alpha} - b\boldsymbol{g}^T \boldsymbol{\Sigma}^{-1} \boldsymbol{g}. \end{aligned}$$

Setting $b = 0$, $\boldsymbol{\Sigma} = \hat{\boldsymbol{\Sigma}}$ and $\boldsymbol{\alpha} = \hat{\boldsymbol{\alpha}}$ gives

$$\boldsymbol{g}^T \hat{\boldsymbol{\Sigma}}^{-1} \boldsymbol{y} - \boldsymbol{g}^T \hat{\boldsymbol{\Sigma}}^{-1} \boldsymbol{X}(\boldsymbol{X}^T \hat{\boldsymbol{\Sigma}}^{-1} \boldsymbol{X})^{-1} \boldsymbol{X}^T \hat{\boldsymbol{\Sigma}}^{-1} \boldsymbol{y} = \boldsymbol{g}^T \boldsymbol{P} \boldsymbol{y},$$

as required. Taking second derivates gives the information matrix:

$$\boldsymbol{\mathcal{I}}(\boldsymbol{\alpha}, b) = \begin{pmatrix} \boldsymbol{g}^T \boldsymbol{\Sigma}^{-1} \boldsymbol{g} & \boldsymbol{g}^T \boldsymbol{\Sigma}^{-1} \boldsymbol{X} \\ \boldsymbol{X}^T \boldsymbol{\Sigma}^{-1} \boldsymbol{g} & \boldsymbol{X}^T \boldsymbol{\Sigma}^{-1} \boldsymbol{X} \end{pmatrix}.$$

Define $\boldsymbol{A}_{i,j}$ to be the element at the $i$th row and $j$th column of the matrix $\boldsymbol{A}$. Then using the block matrix inverse identity gives us that:

$$V = \boldsymbol{\mathcal{I}}_{1,1}^{-1} = \boldsymbol{g}^T \boldsymbol{\Sigma}^{-1} \boldsymbol{g} - \boldsymbol{g}^T \boldsymbol{\Sigma}^{-1} \boldsymbol{X}^T (\boldsymbol{X}^T \boldsymbol{\Sigma}^{-1} \boldsymbol{X})^{-1} \boldsymbol{X} \boldsymbol{\Sigma}^{-1} \boldsymbol{g} = \boldsymbol{g}^T \boldsymbol{P} \boldsymbol{g}.$$

##### Generalised linear mixed model

Under the generalised linear mixed model we have that

$$U = \frac{\partial ql(\boldsymbol{\alpha}, b, \sigma^2)}{\partial b} = \boldsymbol{g}^T(\boldsymbol{y} - \hat{\boldsymbol{y}}),$$

as derived in the derivation of the GLMM fitting algorithm. In the GLMM the information matrix is

$$\boldsymbol{\mathcal{I}}(\boldsymbol{\alpha}, b, \boldsymbol{u}) = \begin{pmatrix} \boldsymbol{X}^T\boldsymbol{W}\boldsymbol{X} & \boldsymbol{X}^T\boldsymbol{W}\boldsymbol{g} & \boldsymbol{X}^T\boldsymbol{W} \\ \boldsymbol{g}^T\boldsymbol{X}\boldsymbol{W} & \boldsymbol{g}^T\boldsymbol{W}\boldsymbol{g} & \boldsymbol{g}^T\boldsymbol{W} \\ \boldsymbol{W}\boldsymbol{X} & \boldsymbol{W}\boldsymbol{g} & \boldsymbol{W} + \boldsymbol{R}^{-1} \end{pmatrix}.$$

Using the block matrix inverse identity gives

$$V = (\boldsymbol{\mathcal{I}}(\boldsymbol{\alpha}, b, \boldsymbol{u})^{-1})_{2,2} = \boldsymbol{g}^T\boldsymbol{P}\boldsymbol{g}$$

where $\boldsymbol{P} = \boldsymbol{\Sigma}^{-1} - \boldsymbol{\Sigma}^{-1}\boldsymbol{X}(\boldsymbol{X}^T\boldsymbol{\Sigma}^{-1}\boldsymbol{X})^{-1}\boldsymbol{X}^T\boldsymbol{\Sigma}^{-1}$, $\boldsymbol{\Sigma} = \boldsymbol{W}^{-1} + \boldsymbol{R}$.

#### 5 Approximating the mixed model score test variance

As described above, for both the LMM and GLMM models calculating the score test variance, $V$, requires calculating

$$\boldsymbol{g}^T\boldsymbol{P}\boldsymbol{g},$$

where $\boldsymbol{g}$ is the vector of genotypes and $\boldsymbol{P} = \boldsymbol{\Sigma}^{-1} - \boldsymbol{\Sigma}^{-1}\boldsymbol{X}(\boldsymbol{X}^T\boldsymbol{\Sigma}^{-1}\boldsymbol{X})^{-1}\boldsymbol{X}^T\boldsymbol{\Sigma}^{-1}$ is an $n \times n$ matrix arising in the mixed model fitting procedure. This operation has computational complexity $O(n^2)$, where $n$ is the number of samples, and is therefore computationally prohibitive to perform for every variant. To avoid this problem, various approaches for approximating the score-test variance are used. This approaches focus on approximating the score test variance as

$$\boldsymbol{g}^T\boldsymbol{P}\boldsymbol{g} = \alpha\boldsymbol{g}^T\boldsymbol{g},$$

where the constant $\alpha$ is chosen to make the approximation as close as possible. The most commonly used approach for choosing $\alpha$ is the variance ratio approach, first proposed in GRAMMAR-GAMMA and also used in the SAIGE, SAIGE-QTL and BOLT-LMM methods [9, 10, 12, 13]. In this approach we first estimate a variance ratio $r$ for each phenotype as,

$$\hat{r} = \frac{1}{m}\sum_{i=1}^{m} \frac{\boldsymbol{g}_i^T\boldsymbol{P}\boldsymbol{g}_i}{\boldsymbol{g}_i^T\boldsymbol{W}\boldsymbol{g}_i},$$

where $g$ is the genotype vector, $\boldsymbol{P}$ is as above, and $\boldsymbol{W}$ is the working weight matrix from the IRWLS algorithm. (For the LMM $\boldsymbol{W} = \boldsymbol{I}$ Usually, $m$ is taken to be small $\approx 30$ and the variants are drawn at random genome-wide.) Then we set $\alpha = r$ to approximate the score test variance as

$$\boldsymbol{g}^T\boldsymbol{P}\boldsymbol{g} \approx \hat{r}\boldsymbol{g}^T\boldsymbol{W}\boldsymbol{g},$$

noting that computing $\boldsymbol{g}^T\boldsymbol{W}\boldsymbol{g}$ is $O(n)$. It has been shown in linear, logistic and Poisson mixed models that $\hat{r}$ is approximately constant across variants [10]. Various versions of the general approach have been implemented, such as the two-stage approximation implemented in SAIGE-QTL.

An alternate approach is used in regenie, a widely used GWAS method [14]. Unlike other methods, regenie uses regularised regressions rather than mixed models to account for population structure across individuals. The regularised regression predictions, $\hat{\boldsymbol{y}}$, are used to form corrected phenotype values $\tilde{\boldsymbol{y}} = \boldsymbol{y} - \hat{\boldsymbol{y}}$, in the same way as in the first step of the overall quasar algorithm. Then in the per-variant score test regenie sets the variance $V$ as

$$V = \hat{\sigma}^2 \boldsymbol{g}^T \boldsymbol{W} \boldsymbol{g} \quad \text{where} \quad \hat{\sigma}^2 = \frac{\tilde{\boldsymbol{y}}^T \tilde{\boldsymbol{y}}}{n - c},$$

where $c$ is the number of covariates. This strategy sets $\alpha$ to the residual variance of each phenotype, removing the the need for the computationally expensive estimation of $r$ for every phenotype, but the reason for its effectiveness is unclear. The paper describing regenie only notes that '[the regenie authors] found in applications that the results obtained using this simple form match up closely to those using a [variance ratio] calibration factor'.

#### Novel trace-based approximation

Here, we introduce our new analytic trance-based approximation. Let $\boldsymbol{A}$ be an $n \times n$ symmetric matrix. Recall that for a $n \times 1$ random vector $\boldsymbol{x}$ with mean $\boldsymbol{\mu}$ and covariance $\boldsymbol{\Sigma}$,

$$\mathbf{E}(\boldsymbol{x}^T \boldsymbol{A} \boldsymbol{x}) = \boldsymbol{\mu}^T \boldsymbol{A} \boldsymbol{\mu} + \operatorname{tr}(\boldsymbol{A} \boldsymbol{\Sigma}).$$

First, we consider the case of a LMM where we have that

$$\hat{r} = \frac{1}{m} \sum_{i=1}^{m} \frac{\tilde{\boldsymbol{g}}^T \boldsymbol{P} \tilde{\boldsymbol{g}}}{\tilde{\boldsymbol{g}}^T \tilde{\boldsymbol{g}}},$$

where $\boldsymbol{P} = \hat{\boldsymbol{\Sigma}}^{-1} - \hat{\boldsymbol{\Sigma}}^{-1} \boldsymbol{X} (\boldsymbol{X}^T \hat{\boldsymbol{\Sigma}}^{-1} \boldsymbol{X})^{-1} \boldsymbol{X}^T \hat{\boldsymbol{\Sigma}}^{-1}$ and $\tilde{\boldsymbol{g}} = \boldsymbol{P}_{\boldsymbol{X}}$, where $\boldsymbol{P}_{\boldsymbol{X}} = \boldsymbol{I} - \boldsymbol{X}(\boldsymbol{X}^T \boldsymbol{X})^{-1} \boldsymbol{X}^T$. $\boldsymbol{P}_{\boldsymbol{X}}$ is a projection matrix, so $\boldsymbol{P}_{\boldsymbol{X}}^2 = \boldsymbol{P}_{\boldsymbol{X}}$. Also, direct calculation shows that $\boldsymbol{P} \boldsymbol{P}_{\boldsymbol{X}} = \boldsymbol{P}$.

We seek to compute the expectation of $\hat{r}$. We assume that $\tilde{\boldsymbol{g}} \sim N(\boldsymbol{0}, \boldsymbol{P}_{\boldsymbol{X}})$. This occurs exactly if $\boldsymbol{g} \sim N(\boldsymbol{0}, \boldsymbol{I})$, but could also occur after applying the residualisation operator $\boldsymbol{P}_{\boldsymbol{X}}$ to non-normally distributed $\boldsymbol{g}$. Then

$$\mathbf{E}(\hat{r}) = \frac{1}{m} \sum_{i=1}^{m} \mathbf{E}\left( \frac{\tilde{\boldsymbol{g}}^T \boldsymbol{P} \tilde{\boldsymbol{g}}}{\tilde{\boldsymbol{g}}^T \tilde{\boldsymbol{g}}} \right) \approx \frac{1}{m} \sum_{i=1}^{m} \frac{\mathbf{E}(\tilde{\boldsymbol{g}}^T \boldsymbol{P} \tilde{\boldsymbol{g}})}{\mathbf{E}(\tilde{\boldsymbol{g}}^T \tilde{\boldsymbol{g}})} = \frac{1}{m} \sum_{i=1}^{m} \frac{\operatorname{tr}(\boldsymbol{P} \boldsymbol{P}_{\boldsymbol{X}})}{\operatorname{tr}(\boldsymbol{P}_{\boldsymbol{X}})} = \frac{1}{m} \sum_{i=1}^{m} \frac{\operatorname{tr}(\boldsymbol{P})}{n - c} = \frac{\operatorname{tr}(\boldsymbol{P})}{n - c}.$$

In this derivation we used a (first order) Taylor series approximation to approximate

$$\mathbf{E}\left( \frac{X}{Y} \right) \approx \frac{\mathbf{E}(X)}{\mathbf{E}(Y)},$$

and that $\operatorname{tr}(\boldsymbol{P}_{\boldsymbol{X}}) = \operatorname{tr}(\boldsymbol{I} - \boldsymbol{X}(\boldsymbol{X}^T \boldsymbol{X})^{-1} \boldsymbol{X}^T) = n - c$, where $n$ is the number of rows of $\boldsymbol{X}$ and $c$ is the number of columns. In simulations the agreement between the variance-ratio approximation and the trace-based approximation is almost perfect (Figure 1a). Second, in the case of the GLMM we have that

$$\hat{r} = \frac{1}{m} \sum_{i=1}^{m} \frac{\tilde{\boldsymbol{g}} \boldsymbol{P} \tilde{\boldsymbol{g}}}{\tilde{\boldsymbol{g}}^T \boldsymbol{W} \tilde{\boldsymbol{g}}},$$

where now $\tilde{\boldsymbol{g}} = \boldsymbol{P}_{\boldsymbol{W}} \boldsymbol{g}$ where $\boldsymbol{P}_{\boldsymbol{W}} = \boldsymbol{I} - \boldsymbol{X}(\boldsymbol{X}^T \boldsymbol{W} \boldsymbol{X})^{-1} \boldsymbol{X} \boldsymbol{W}$. As for $\boldsymbol{P}_{\boldsymbol{X}}$ it is true that $\boldsymbol{P} \boldsymbol{P}_{\boldsymbol{W}} = \boldsymbol{P}$. Using the same argument as above, and noting that

$$\mathbf{E}(\tilde{\boldsymbol{g}}^T \boldsymbol{W} \tilde{\boldsymbol{g}}) = \operatorname{tr}(\boldsymbol{W} \boldsymbol{P}_{\boldsymbol{W}}),$$

we can compute

$$\mathbf{E}(\hat{r}) = \frac{\text{tr}(\boldsymbol{P})}{\text{tr}(\boldsymbol{W}\boldsymbol{P}_{\boldsymbol{W}})}.$$

However in practice we found that this approximation did not perform well in simulations, likely due to not accounting for the correlation of the numerator and denominator as $\boldsymbol{P}$ depends on $\boldsymbol{W}$ through $\boldsymbol{\Sigma}^{-1}$ (Figure 1c). We therefore use the second order Taylor expansion for the expectation of the ratio

$$\mathbf{E}\left(\frac{X}{Y}\right) \approx \frac{\mathbf{E}(X)}{\mathbf{E}(Y)} - \frac{\text{Cov}(X,Y)}{\mathbf{E}(X)^2} + \frac{\text{Var}(X)\mathbf{E}(Y)}{\mathbf{E}(X)^3}.$$

We use the following expressions for the variance and covariance of a quadratic form (which rely on the additional assumption that the random vector follows a Gaussian distribution). If $\boldsymbol{x} \sim N(\boldsymbol{\mu}, \boldsymbol{\Sigma})$ and $\boldsymbol{A}$ and $\boldsymbol{B}$ are symmetric matrices, then

$$\text{Var}(\boldsymbol{x}^T\boldsymbol{A}\boldsymbol{x}) = 2\text{tr}(\boldsymbol{A}\boldsymbol{\Sigma}\boldsymbol{A}\boldsymbol{\Sigma}) + 4\boldsymbol{\mu}^T\boldsymbol{A}\boldsymbol{\Sigma}\boldsymbol{A}\boldsymbol{\Sigma}\boldsymbol{\mu}$$

and

$$\text{Cov}(\boldsymbol{x}^T\boldsymbol{A}\boldsymbol{x}, \boldsymbol{x}^T\boldsymbol{B}\boldsymbol{x}) = 2\text{tr}(\boldsymbol{A}\boldsymbol{\Sigma}\boldsymbol{B}\boldsymbol{\Sigma}) + 4\boldsymbol{\mu}^T\boldsymbol{A}\boldsymbol{\Sigma}\boldsymbol{B}\boldsymbol{\Sigma}\boldsymbol{\mu}.$$

Combining these expression leads to the approximate expectation

$$\mathbf{E}(\hat{r}) \approx \frac{\text{tr}(\boldsymbol{P})}{\text{tr}(\boldsymbol{W}\boldsymbol{P}_{\boldsymbol{W}})} - \frac{2\text{tr}(\boldsymbol{P}\boldsymbol{W}\boldsymbol{P}_{\boldsymbol{W}})}{\text{tr}(\boldsymbol{W}\boldsymbol{P}_{\boldsymbol{W}})^2} + \frac{2\text{tr}(\boldsymbol{W}\boldsymbol{P}_{\boldsymbol{W}}\boldsymbol{W}\boldsymbol{P}_{\boldsymbol{W}})\text{tr}(\boldsymbol{P})}{\text{tr}(\boldsymbol{W}\boldsymbol{P}_{\boldsymbol{W}})^3}.$$

This second order approximation performs much better than the first order approximation in simulations, though the approximation is not as good as in the LMM case (Figure 1d). Unlike the LMM estimator which can be computed in $O(n)$ time, the second order estimator requires matrix multiplication. In quasar, we use properties of the trace and pre-computation of key matrices to optimize the calculation of the approximation. The computation's most expensive operation is the multiplication of the $n \times n$ matrix $\boldsymbol{P}$ with the $n \times c$ matrix $\boldsymbol{W}\boldsymbol{X}$ which has computational complexity $O(n^2c)$. Computing the variance-ratio approximation has complexity $O(n^2m)$ where $m$ is the number of variants summed over. Therefore we expect the second-order trace-based approximation to have similar performance to the variance-ratio approximation, and likely better as generally $c$, the number of covariates is less than $m$, which is at least 30 in most implementations of the variance-ratio approximation.

##### Connection to regenie approximation

Our estimator is closely connected to the one used in regenie. Consider the linear mixed model

$$\boldsymbol{y} = \boldsymbol{X}\boldsymbol{\alpha} + \boldsymbol{g}b + \boldsymbol{e}, \quad \boldsymbol{u} \sim N(\mathbf{0}, \boldsymbol{\Sigma}), \text{ where } \boldsymbol{\Sigma} = \sigma_g^2\boldsymbol{K} + \sigma_e^2\boldsymbol{I}.$$

We can fit the model under the null $b = 0$ to estimate $\sigma_g^2$ and $\sigma_e^2$, therefore estimating $\boldsymbol{\Sigma}$. We form the residualisation matrix $\boldsymbol{P} = \boldsymbol{\Sigma}^{-1} - \boldsymbol{\Sigma}^{-1}\boldsymbol{X}(\boldsymbol{X}^T\boldsymbol{\Sigma}^{-1}\boldsymbol{X})^{-1}\boldsymbol{X}^T\boldsymbol{\Sigma}^{-1}$ which we used to construct $\tilde{\boldsymbol{y}} = \boldsymbol{P}\boldsymbol{y}$ for use in the score test. We can compute the distribution of $\tilde{\boldsymbol{y}}$ under the null. As the linear transformation of a normal distribution, $\tilde{\boldsymbol{y}}$ will have a normal distribution. We first derive a useful identity, that $\boldsymbol{P\Sigma P} = \boldsymbol{P}$:

$$\begin{aligned}
\boldsymbol{P\Sigma P} &= (\boldsymbol{\Sigma}^{-1} - \boldsymbol{\Sigma}^{-1}\boldsymbol{X}(\boldsymbol{X}^T\boldsymbol{\Sigma}^{-1}\boldsymbol{X})^{-1}\boldsymbol{X}^T\boldsymbol{\Sigma}^{-1})\boldsymbol{\Sigma}(\boldsymbol{\Sigma}^{-1} - \boldsymbol{\Sigma}^{-1}\boldsymbol{X}(\boldsymbol{X}^T\boldsymbol{\Sigma}^{-1}\boldsymbol{X})^{-1}\boldsymbol{X}^T\boldsymbol{\Sigma}^{-1} \\
&= \boldsymbol{\Sigma}^{-1} - \boldsymbol{\Sigma}^{-1}\boldsymbol{X}(\boldsymbol{X}^T\boldsymbol{\Sigma}^{-1}\boldsymbol{X})^{-1}\boldsymbol{X}^T\boldsymbol{\Sigma}^{-1} - \boldsymbol{\Sigma}^{-1}\boldsymbol{X}(\boldsymbol{X}^T\boldsymbol{\Sigma}^{-1}\boldsymbol{X})^{-1}\boldsymbol{X}^T\boldsymbol{\Sigma}^{-1} \\
&\quad + \boldsymbol{\Sigma}^{-1}\boldsymbol{X}(\boldsymbol{X}^T\boldsymbol{\Sigma}^{-1}\boldsymbol{X})^{-1}\boldsymbol{X}^T\boldsymbol{\Sigma}^{-1}\boldsymbol{X}(\boldsymbol{X}^T\boldsymbol{\Sigma}^{-1}\boldsymbol{X})^{-1}\boldsymbol{X}^T\boldsymbol{\Sigma}^{-1} \\
&= \boldsymbol{\Sigma}^{-1} - \boldsymbol{\Sigma}^{-1}\boldsymbol{X}(\boldsymbol{X}^T\boldsymbol{\Sigma}^{-1}\boldsymbol{X})^{-1}\boldsymbol{X}^T\boldsymbol{\Sigma}^{-1} \\
&= \boldsymbol{P}.
\end{aligned}$$

First,

$$\mathbf{E}(\tilde{\boldsymbol{y}}) = \mathbf{E}(\boldsymbol{P y}) = \boldsymbol{P}\mathbf{E}(\boldsymbol{y}) = \boldsymbol{P X \alpha}.$$

Evaluating this expression at the estimate of $\boldsymbol{\alpha}$: $\hat{\boldsymbol{\alpha}} = \boldsymbol{\Sigma}^{-1}\boldsymbol{X}(\boldsymbol{X}^T\boldsymbol{\Sigma}^{-1}\boldsymbol{X})^{-1}\boldsymbol{X}^T\boldsymbol{\Sigma}^{-1}\boldsymbol{y} = (\boldsymbol{I} - \boldsymbol{\Sigma P})\boldsymbol{y}$, gives

$$\mathbf{E}(\tilde{\boldsymbol{y}}) = \boldsymbol{P X}\hat{\boldsymbol{\alpha}} = \boldsymbol{P}(\boldsymbol{I} - \boldsymbol{\Sigma P})\boldsymbol{y} = (\boldsymbol{P} - \boldsymbol{P\Sigma P})\boldsymbol{y} = (\boldsymbol{P} - \boldsymbol{P})\boldsymbol{y} = \boldsymbol{0},$$

where we used the identify $\boldsymbol{P\Sigma P} = \boldsymbol{P}$. We have that $\tilde{\boldsymbol{y}} = \boldsymbol{P y} \sim N(\boldsymbol{0}, \boldsymbol{P})$. Second, we have that

$$\text{Var}(\tilde{\boldsymbol{y}}) = \text{Var}(\boldsymbol{P y}) = \boldsymbol{P\Sigma P} = \boldsymbol{P}.$$

Overall, $\tilde{\boldsymbol{y}} \sim N(\boldsymbol{0}, \boldsymbol{P})$, Finally, recalling the expectation of a quadratic form, we have

$$\mathbf{E}(\tilde{\boldsymbol{y}}^T\tilde{\boldsymbol{y}}) = \text{tr}(\boldsymbol{P})$$

Therefore, under a linear mixed model, the expected value of the regenie approximation is equal to our novel approximation, that is,

$$\mathbf{E}\left(\frac{\tilde{\boldsymbol{y}}^T\tilde{\boldsymbol{y}}}{n-c}\right) = \frac{\text{tr}(\boldsymbol{P})}{n-c}.$$

In simulations the concordance between the trace-based and regenie approximations is very good (Figure 1b). While regenie doesn't use a linear mixed model to correct for sample relatedness, this derivation highlights that its approximation can also be viewed as estimating the trace of the residualisation matrix $\boldsymbol{P}$. In addition, this derivation shows that in expectation both the variance-ratio approximation and regenie's approximation equal the approximation introduced in this paper.

#### Simulations

To assess the performance of the approximations we performed a small simulation study. Briefly, we set the the number of individuals to be 100, and used the top right corner of the OneK1K computed GRM as the random effect. We simulate genotypes across 100 SNPs as Bi(2, 0.2). We simulate both an LMM and Poisson GLMM model. The null LMM is fit using the `mixed.solve()` function from the `rrBLUP` package. The null GLMM is fit using `lme4` package. The variance ratio approximation is calculated across all SNPs.

#### Covariate-adjusted genotypes

In practice, in the score test formulas we replace the genotype vector $\boldsymbol{g}$ by the 'residualised' covariate adjusted genotype vector, $\tilde{\boldsymbol{g}}$, defined by

$$\tilde{\boldsymbol{g}} = \boldsymbol{P_W g} = \boldsymbol{g} - \boldsymbol{X}(\boldsymbol{X}^T\boldsymbol{W X})^{-1}\boldsymbol{X}^T\boldsymbol{W g},$$

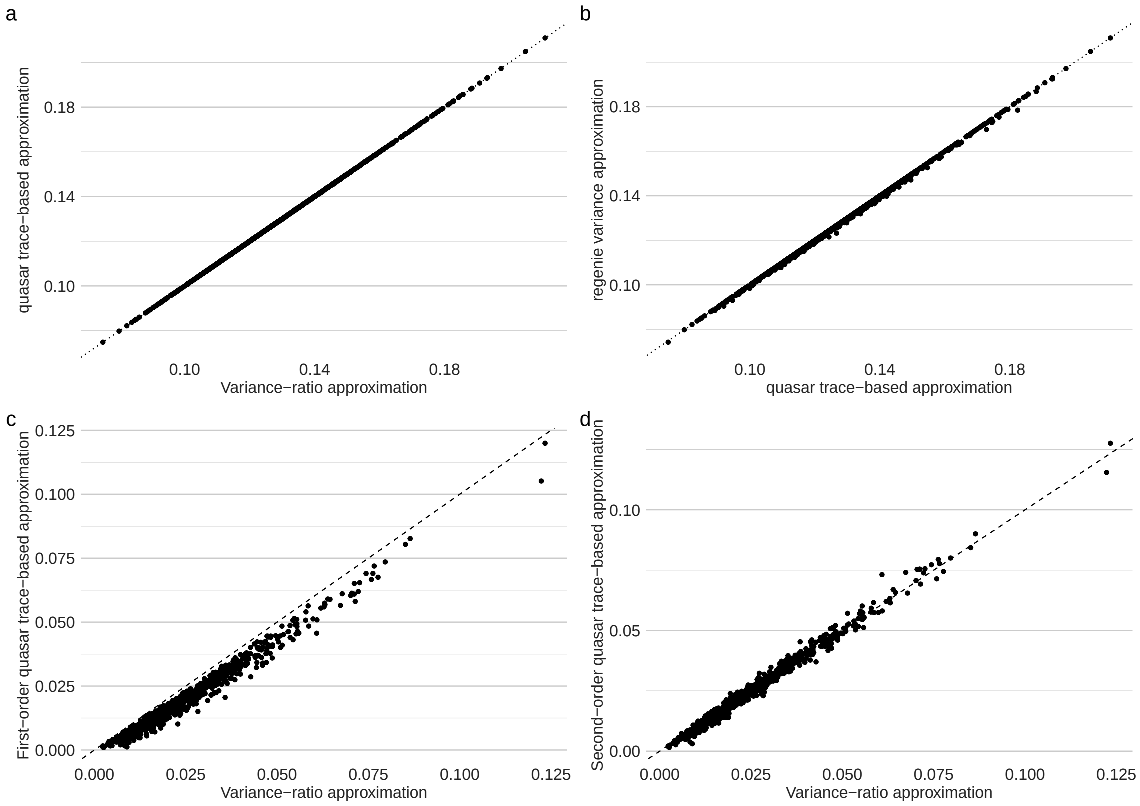


Figure 1: **Performance of trace-base approximation in simulations** a) LMM case, trace-based approximation plotted against the variance-ratio approximation b) LMM case, trace-based approximation plotted against regenie approximation. c) GLMM case, first-order trace-based approximation plotted against the variance-ratio approximation. d) GLMM case, second-order trace-based approximation plotted against the variance-ratio approximation

where $\boldsymbol{X}$ is the matrix of covariates and $\boldsymbol{W}$ is the estimated working-weight matrix. (In the LM and LMM $\boldsymbol{W} = \boldsymbol{I}$). Although the covariate adjusted genotype vector does not arise from a standard derivation of the score statistic in the context of LMM or GLMM models it is true that

$$\tilde{\boldsymbol{g}}^T \boldsymbol{P} \tilde{\boldsymbol{g}} = (\boldsymbol{P}_{\boldsymbol{W}} \boldsymbol{g})^T \boldsymbol{P} (\boldsymbol{P}_{\boldsymbol{W}} \boldsymbol{g}) = \boldsymbol{g}^T \boldsymbol{P}_{\boldsymbol{W}}^T \boldsymbol{P} \boldsymbol{P}_{\boldsymbol{W}} \boldsymbol{g} = \boldsymbol{g}^T \boldsymbol{P} \boldsymbol{g},$$

and the covariate adjusted score vector arises in the derivation of the score test in the GLM/LM case [15].

### Multiple hypothesis correction

Following other methods for QTL mapping we use a two-stage approach to performing multiple testing correction.

First, for each gene the aggregated Cauchy association test (ACAT) is used to provide an aggregated summary of the significance of the gene [16]. The ACAT was first used for this purpose in APEX and is also used by SAIGE-QTL [6, 10]. Prior to the use of the ACAT, a permutation approach was used [17], however, repeatedly performing the QTL mapping is computationally expensive. For a vector of p-values $p_1, \ldots, p_k$ the ACAT statistic is defined as

$$T_{\mathrm{ACAT}} = \sum_{i=1}^{k} \tan\left(\left(\frac{1}{2} - p_i\right)\pi\right),$$

where $\tan(\cdot)$ is the standard tan function. If $p_i \sim U(0,1)$, i.e., under $H_0$, then $\tan((\frac{1}{2} - p_i)\pi)$ follows a standard Cauchy distribution. Under $H_0$, the distribution of $T_{\mathrm{ACAT}}$ is well approximated by a Cauchy distribution with location parameter 0 and scale parameter $k$. Therefore, the associated p-value of the $T_{\mathrm{ACAT}}$, $p_{\mathrm{ACAT}}$ is approximately

$$p_{\mathrm{ACAT}} \approx \frac{1}{2} - \frac{1}{\pi}\arctan\left(\frac{T_{\mathrm{ACAT}}}{k}\right).$$

As all calculations involved in the ACAT involve sums and analytic functions they are computationally inexpensive.

Second, at the level of genes conventionally the q-value method has been used [17]. This method has been used instead of other methods for controlling the FDR as it estimates the probability of null cases, rather than assuming it is one, which is not a plausible assumption in this context as we expect that a high proportion of genes have their expression genetically regulated. However, as discussed in the main text the q-value estimation procedure makes an assumption not satisfied by p-values computed with ACAT so we recommend the classical Benjamini-Hochberg procedure for controlling the FDR when ACAT is used.

[9] Wei Zhou, Jonas B. Nielsen, Lars G. Fritsche, Rounak Dey, Maiken E. Gabrielsen, Brooke N. Wolford, Jonathon LeFaive, Peter VandeHaar, Sarah A. Gagliano, Aliya Gifford, Lisa A. Bastarache, Wei-Qi Wei, Joshua C. Denny, Maoxuan Lin, Kristian Hveem, Hyun Min Kang, Goncalo R. Abecasis, Cristen J. Willer, and Seunggeun Lee. Efficiently controlling for case-control imbalance and sample relatedness in large-scale genetic association studies. *Nature Genetics*, 50(9):1335–1341, September 2018. ISSN 1546-1718. doi: 10.1038/s41588-018-0184-y. URL https://www.nature.com/articles/s41588-018-0184-y. Publisher: Nature Publishing Group.

[10] Wei Zhou, Anna S. E. Cuomo, Angli Xue, Masahiro Kanai, Grant Chau, Chirag Krishna, Ramnik J. Xavier, Daniel G. MacArthur, Joseph E. Powell, Mark J. Daly, and Benjamin M. Neale. Efficient and accurate mixed model association tool for single-cell eQTL analysis, May 2024. URL https://www.medrxiv.org/content/10.1101/2024.05.15.24307317v1. Pages: 2024.05.15.24307317.

[11] Peter K. Dunn and Gordon Smyth. *Generalized Linear Models With Examples in R*. Springer New York, NY, New York, 2018.

[12] Gulnara R. Svishcheva, Tatiana I. Axenovich, Nadezhda M. Belonogova, Cornelia M. van Duijn, and Yurii S. Aulchenko. Rapid variance components–based method for whole-genome association analysis. *Nature Genetics*, 44(10):1166–1170, October 2012. ISSN 1546-1718. doi: 10.1038/ng.2410. URL https://www.nature.com/articles/ng.2410. Publisher: Nature Publishing Group.

[13] Po-Ru Loh, George Tucker, Brendan K. Bulik-Sullivan, Bjarni J. Vilhjálmsson, Hilary K. Finucane, Rany M. Salem, Daniel I. Chasman, Paul M. Ridker, Benjamin M. Neale, Bonnie Berger, Nick Patterson, and Alkes L. Price. Efficient Bayesian mixed-model analysis increases association power in large cohorts. *Nature Genetics*, 47(3):284–290, March 2015. ISSN 1546-1718. doi: 10.1038/ng.3190. URL https://www.nature.com/articles/ng.3190. Publisher: Nature Publishing Group.

[14] Joelle Mbatchou, Leland Barnard, Joshua Backman, Anthony Marcketta, Jack A. Kosmicki, Andrey Ziyatdinov, Christian Benner, Colm O'Dushlaine, Mathew Barber, Boris Boutkov, Lukas

Habegger, Manuel Ferreira, Aris Baras, Jeffrey Reid, Goncalo Abecasis, Evan Maxwell, and Jonathan Marchini. Computationally efficient whole-genome regression for quantitative and binary traits. *Nature Genetics*, 53(7):1097–1103, July 2021. ISSN 1546-1718. doi: 10.1038/s41588-021-00870-7. URL https://www.nature.com/articles/s41588-021-00870-7. Publisher: Nature Publishing Group.

[15] Rounak Dey, Ellen M. Schmidt, Goncalo R. Abecasis, and Seunggeun Lee. A Fast and Accurate Algorithm to Test for Binary Phenotypes and Its Application to PheWAS. *The American Journal of Human Genetics*, 101(1):37–49, July 2017. ISSN 0002-9297, 1537-6605. doi: 10.1016/j.ajhg.2017.05.014. URL https://www.cell.com/ajhg/abstract/S0002-9297(17)30201-X. Publisher: Elsevier.

[16] Yaowu Liu and Jun Xie. Cauchy Combination Test: A Powerful Test With Analytic $p$-Value Calculation Under Arbitrary Dependency Structures. *Journal of the American Statistical Association*, 115(529):393–402, January 2020. ISSN 0162-1459, 1537-274X. doi: 10.1080/01621459.2018.1554485. URL https://www.tandfonline.com/doi/full/10.1080/01621459.2018.1554485.

[17] Halit Ongen, Alfonso Buil, Andrew Anand Brown, Emmanouil T. Dermitzakis, and Olivier Delaneau. Fast and efficient QTL mapper for thousands of molecular phenotypes. *Bioinformatics*, 32(10):1479–1485, May 2016. ISSN 1367-4803. doi: 10.1093/bioinformatics/btv722. URL https://doi.org/10.1093/bioinformatics/btv722.
